# Persistent PTSD symptoms are associated with plasma metabolic alterations relevant to long-term health: A metabolome-wide investigation in women

**DOI:** 10.1101/2024.08.07.24311628

**Authors:** Yiwen Zhu, Katherine H. Shutta, Tianyi Huang, Raji Balasubramanian, Oana A. Zeleznik, Clary B. Clish, Julián Ávila-Pacheco, Susan E. Hankinson, Laura D. Kubzansky

**Author notes:** Both senior authors contributed equally (Susan E. Hankinson, Laura D. Kubzansky). **Correspondence**: Yiwen Zhu, MS, Department of Epidemiology, Harvard T.H. Chan School of Public Health, 677 Huntington Avenue, Boston, MA 02115.

## Abstract

**Background:** Posttraumatic stress disorder (PTSD) is characterized by severe distress and associated with cardiometabolic diseases. Studies in military and clinical populations suggest dysregulated metabolomic processes may be a key mechanism. Prior work identified and validated a metabolite-based distress score (MDS) linked with depression and anxiety and subsequent cardiometabolic diseases. Here, we assessed whether PTSD shares metabolic alterations with depression and anxiety and also if additional metabolites are related to PTSD.

**Methods:** We leveraged plasma metabolomics data from three subsamples nested within the Nurses’ Health Study II, including 2835 women with 2950 blood samples collected across three timepoints (1996-2014) and 339 known metabolites consistently assayed by mass spectrometry-based techniques. Trauma and PTSD exposures were assessed in 2008 and characterized as follows: lifetime trauma without PTSD, lifetime PTSD in remission, and persistent PTSD symptoms. Associations between the exposures and the MDS or individual metabolites were estimated within each subsample adjusting for potential confounders and combined in random-effects meta-analyses.

**Results:** Persistent PTSD symptoms were associated with higher levels of the previously developed MDS for depression and anxiety. Out of 339 metabolites, we identified nine metabolites (primarily elevated glycerophospholipids) associated with persistent symptoms (false discovery rate<0.05). No metabolite associations were found with the other PTSD-related exposures.

**Conclusions:** As the first large-scale, population-based metabolomics analysis of PTSD, our study highlighted shared and distinct metabolic differences linked to PTSD versus depression or anxiety. We identified novel metabolite markers associated with PTSD symptom persistence, suggesting further connections with metabolic dysregulation that may have downstream consequences for health.

## Introduction

Posttraumatic stress disorder (PTSD) is a debilitating psychiatric disorder characterized by severe distress and occurs after experiencing or witnessing a traumatic event. Lifetime prevalence of PTSD is estimated to be 7-8% among US adults, and women are more than twice as likely to develop PTSD compared to men (Kilpatrick et al., 2013). PTSD has also been linked with adverse physical health, including higher inflammation (Sumner et al., 2020), elevated cardiometabolic risks (Sumner et al., 2015), higher incidence of certain cancers, and early mortality (Roberts et al., 2019, 2020). Experimental studies support overlap in PTSD-related pathophysiology and mechanisms contributing to the susceptibility and progression of cardiometabolic diseases (Miller et al., 2011). As such, PTSD is not only a brain disorder, but also a systemic illness with a wide range of physical manifestations (Michopoulos et al., 2016; Mellon et al., 2018), including metabolic alterations.

Metabolites are small molecules that can be metabolized and occur in circulation, such as lipids or amino acids (Wishart et al., 2022). They are considered the endpoint along the “omics cascade” most proximal to behaviors and diseases, capturing additional information beyond upstream genetic and transcriptomic regulations (Clish, 2015; Patti et al., 2012). Studies on PTSD and metabolites have mostly focused on *a priori* selected candidates, such as steroids and steroid derivatives implicated in glucocorticoid metabolism, yielding null or inconsistent findings (Zhu et al., 2022). Another approach to metabolomic analyses is an agnostic investigation of all quantified metabolites in a biological system or specimen (Dunn et al., 2005). To our knowledge, four agnostic metabolomic studies of PTSD to date (Karabatsiakis et al., 2015; Mellon et al., 2019; Konjevod et al., 2021; Kuan et al., 2022) suggest that alterations of mitochondrial functioning, fatty acid metabolism, and lipid metabolism may be involved in PTSD pathophysiology. However, compared with metabolomic studies of other psychological conditions (e.g., depression, Bot et al., 2020; Davyson et al., 2023), these studies were relatively small (N=38 to 204), with limited power to detect signals of individual metabolites, despite intriguing patterns revealed in pathway-or module-level analyses. Further, three included only male participants; no metabolomic analyses of PTSD have been performed in population-based cohorts of civilian women. Whether metabolic alterations persist following PTSD remission also remains poorly understood.

Metabolomic studies of other forms of psychological distress, such as depression and anxiety, have established linkages between distress and metabolic alterations related to cardiometabolic health (Huang et al., 2020; Pu et al., 2020; van der Spek et al., 2023). Earlier work from our group identified and validated a metabolomic signature for chronic depression or anxiety in two cohorts of women (Shutta et al., 2021). We subsequently developed a metabolite-based distress score (MDS) comprised of 20 plasma metabolites. Notably, this MDS was associated with increased risk of incident cardiovascular disease (CVD) and diabetes in independent samples (Balasubramanian et al., 2023; Huang et al., 2023). Given PTSD is highly co-morbid and shares similar symptoms with both anxiety and depression and has also been reliably associated with higher risk of CVD and diabetes (Galatzer-Levy et al., 2013), it is possible that similar metabolic variations may be observed among individuals with PTSD.

The current study leveraged plasma metabolomics data from three subsamples nested within an ongoing cohort of predominantly civilian women in the Nurses’ Health Study II (NHSII) to address two primary research questions. First, we assessed whether PTSD shares common metabolic alterations with depression and anxiety by testing associations of PTSD with the previously derived MDS and its individual components (Balasubramanian et al., 2023). Of note, there was no overlap between the sample used to develop the MDS and the analytic sample used for the current study. We hypothesized (a) PTSD would be associated with the MDS and its individual metabolite components; and (b) the associations would be stronger among individuals with active, persistent PTSD symptoms versus with remitted PTSD. Second, to identify other markers related to PTSD symptoms, we conducted an agnostic metabolome-wide investigation of 339 individual metabolites and a differential network analysis characterizing systems-level differences. These provide complementary insights to the agnostic analyses by highlighting patterns of connections between metabolites. To address potential confounding, based on prior literature demonstrating the impact of biomedical and behavioral variables on circulating metabolite concentrations (e.g., Boulet et al., 2015; Fujisaka et al., 2018), our analyses adjusted for demographics, medical history and health conditions, and health-related behaviors.

## Methods and Materials

### Study population

NHSII is an ongoing, prospective cohort of 116,429 female registered nurses residing in 14 US states at enrollment in 1989. Participants were between 25 to 42 years of age at baseline and have completed biennial follow-up surveys for over three decades. Analytic samples for the present study are from three independent subsamples of participants nested within the NHSII cohort, all of whom completed a questionnaire in 2008 that assessed lifetime exposure to traumatic events and PTSD. Consistent data collection, quality control, and preprocessing protocols for both questionnaire-based and metabolomic data have been implemented for all participants. The **first subsample** draws on recent cohort-wide efforts to harmonize metabolomics data across multiple sub-studies of various disease endpoints (hereafter referred to as the *NHSII merged dataset*), from which we identified 2550 samples from 2435 women across two blood collections (2276 in 1996-1999, 44 in 2010-2012 and 115 with blood samples taken at both time points); The **second subsample** draws on 204 women in the Mind-Body Study (*MBS*), an NHSII sub-study designed to investigate connections between psychosocial factors and biological processes (Huang et al., 2019), with metabolomics data obtained from blood collections between 2013-2014. The **third subsample** (hereafter referred to as the *Severe Distress Sample*) draws on new metabolomic data obtained from blood samples collected between 2010-2012 among 196 NHSII participants, oversampling women with significant PTSD exposure. See **Supplement** and **Supplemental Figure S1** for a detailed description of each analytic subsample and a study timeline.

This study was approved by the institutional review board of the Brigham and Women’s Hospital and Harvard T.H. Chan School of Public Health; participants’ completion of questionnaires implied informed consent based on the study protocol.

### PTSD and trauma measures

PTSD and trauma were assessed in 2008 using the same measures for all study subsamples. Lifetime trauma exposure was assessed using the 16-item modified Brief Trauma Questionnaire (Schnurr et al., 2002), capturing experiences of 15 potentially traumatic events (e.g., life threats or serious injury) and “a seriously traumatic event not already covered” based on DSM-IV diagnostic criteria (American Psychiatric Association, 1994). Participants reported their first trauma exposure, worst trauma exposure (if they experienced multiple exposures), and if exposed, their ages at the first and worst trauma events. We created an indicator variable for trauma exposure (exposed/unexposed). Participants were considered unexposed to trauma at a given blood collection if their first trauma occurred after blood collection or if they reported no trauma exposures as of 2008. Participants were prompted to answer questions about PTSD symptoms if they indicated any trauma exposure, following the required diagnostic criteria.

Lifetime and past-month PTSD symptoms were measured with the 7-item Short Screening Scale for DSM-IV PTSD (Breslau et al., 1999), referencing the worst trauma identified by the respondent. For each of seven symptoms, participants indicated (i) whether they ever experienced the symptom and (ii) if they experienced it in the past four weeks. Two PTSD variables were derived: *persistent PTSD symptoms* - the count of PTSD symptoms experienced within the past four weeks at the time of reporting in 2008 (range=0-7); *remitted PTSD-* a binary indicator capturing whether the participant experienced lifetime PTSD (reporting ≥4 symptoms) but did not report experiencing any symptoms in the past-month (Ratanatharathorn et al., 2022; Sumner et al., 2015). The binary cut-point of ≥4 symptoms has been shown to capture PTSD cases with a sensitivity of 85% and a specificity of 93% (Breslau et al., 1999).

### Blood collection and metabolomic profiling

For all blood collections, participants arranged to have their blood samples collected and processed following standard procedures; samples were stored at −130 C° or lower. Across all subsamples, plasma metabolomic profiling was performed at the Broad Institute of the Massachusetts Institute of Technology and Harvard using a platform comprising complementary liquid chromatography tandem mass spectrometry (LC-MS)-based methods. 339 metabolites were available in all three subsamples with missingness <50% within each sub-study sample. Additional technical details about metabolite selection, profiling, and quality control are described elsewhere (Paynter et al., 2018) and in the **Supplement**.

To assess metabolic alterations linked to depression and anxiety we calculated a metabolite-based distress score (MDS), following prior work in a similar cohort of women (Balasubramanian et al., 2023). Of note, 19 out of the 20 metabolites identified in the original publication are available in our analytic subsamples and therefore included in the MDS. See **Supplement** for details of the MDS development and individual metabolite weights.

### Covariate measures

Three sets of covariates are considered incrementally in the analyses. Unless stated otherwise, all covariates were taken from self-report assessments closest in time to the relevant blood collection for each subsample and queried in the same way. (1) Minimal model covariates: demographic variables and matching factors from the sub-studies that followed a case-control design, namely, age, race/ethnicity, menopausal status, fasting status at blood collection, and sub-study indicator (only applicable in the NHSII merged dataset); (2) medical covariates: use of statins or other lipid lowering drugs, hormone therapy, hypertension history, and type 2 diabetes history; (3) biobehavioral covariates: diet quality (measured by the Alternative Healthy Eating Index (Chiuve et al., 2012)), physical activity (measured by the total metabolic equivalent task (MET) hours per week (Ainsworth et al., 1993)), alcohol intake (g/day), smoking status (current/not current), caffeine intake (mg/day), and body-mass index (BMI). See **Supplement** for additional information on covariates.

### Statistical analysis

Metabolite levels were log-transformed and normalized within each lab sample prior to data analysis. For metabolites with <50% missing values, metabolite levels were assumed to be below the detection limit and missing values were imputed as ½ of the minimum observed value. This strategy is consistent with analytic protocols followed by prior cohort publications (Huang et al., 2020; J. Li et al., 2020) and preferred over imputation methods because missingness in metabolomics profiling primarily arises from the true value falling below the limit of detection (i.e., missing not at random, Do et al., 2018).

As missingness in covariates was minimal (up to 4%, for alcohol intake), we performed complete-case analysis. For all analyses, persistent PTSD symptoms, remitted PTSD status, and trauma exposure were included simultaneously in each model as primary predictors for a comprehensive characterization of PTSD (see **Supplement** for coefficient interpretations). Analyses incrementally adjusted for the three sets of covariates described earlier. We accounted for multiple testing by applying false discovery rate (FDR) adjustment to the p-values and presenting results passing the significance threshold of adjusted p<0.05. All analyses were performed using *SAS* version 9.4 and *R* version 4.1.0.

**First,** we assessed whether levels of metabolites previously linked to depression and anxiety were associated with PTSD and trauma exposure. Specifically, we pursued two sets of analyses: (a) testing the association with the MDS (N=2006 with complete data on all 19 metabolites); and (b) testing metabolite-level associations for each of the 19 available metabolites in the MDS. We first performed separate analyses within each subsample using either generalized estimating equations (in the NHSII merged dataset to account for within-individual dependence across the two blood collections) or linear regression (in MBS and the Severe Distress Sample). The three subsample-specific estimates were then combined using a random effects meta-analysis to allow for subsample-specific variations in the exposure effects. Cochran’s Q-statistics were calculated to assess between-study heterogeneity.

**Second,** we performed a metabolome-wide agnostic analysis to identify metabolic markers (that may not have been in the MDS) associated with PTSD and trauma exposures. Extending the metabolite-level analysis described above to all 339 available metabolites, we also meta-analyzed estimates across substudies for the association between each metabolite and PTSD/trauma exposure (N’s ranged from 438 to 2779 per metabolite; 83% of metabolites have N>2000). We primarily present and interpret results obtained from Model 2, which adjusts for both baseline and medical covariates, as they capture key potential confounders. Following the agnostic analysis, to examine metabolite-class level differences between metabolomic associations with PTSD and those with depression/anxiety, we identified metabolite classes enriched for associations with persistent PTSD symptoms from the minimally adjusted meta-analyses and compared with enrichment analysis of depression/anxiety conducted using summary statistics reported in prior research (Shutta et al., 2021).

**Third**, we characterized systems-level metabolic differences linked to PTSD by constructing a differential network between participants who reported any versus no persistent PTSD symptoms (exposed versus unexposed) using the differential network analysis in genomics (DINGO) algorithm (Ha et al., 2015). We constructed our differential network using the 29 metabolites associated with persistent PTSD symptoms in the minimally adjusted analysis. By analyzing the conditional dependencies between metabolite pairs and testing for differences between participants with versus without persistent symptoms, DINGO provides a connected, network-focused view complementary to the agnostic analyses of individual metabolites. In a metabolite network, nodes represent individual metabolites and undirected, weighted edges between node pairs reflect the partial correlation between the corresponding metabolite pair.

Specifically, using DINGO, we weighted the edges by the *difference* in partial correlations of a metabolite pair between participants with persistent symptoms relative to controls. To assess the importance of each metabolite in the network structure, we calculated three different centrality measures: hub centrality (the relative influence of a node in the network) (Kleinberg, 1999), betweenness centrality (the number of shortest paths through the network that include the node (Brandes, 2001; Freeman, 1978)); and closeness centrality (the average distance from each node to all the other nodes in the network (Freeman, 1978)). A node with high values in any or all metrics indicates a metabolite whose patterns of correlations with other metabolites have changed in the presence of persistent symptoms; such node may be a key driver of potential metabolic dysregulations.

See **Supplement** for details on our analytic approaches.

## Results

### Sample characteristics

Participants’ demographic and health characteristics are shown in **Table 1**. Overall, 27.7% of our analytic sample reported persistent PTSD symptoms in 2008, 8% reported no persistent symptoms but met diagnostic criteria for probable lifetime PTSD (i.e., PTSD in remission), 39.7% were trauma exposed but did not have PTSD, and 24.7% were unexposed to trauma. Women reporting persistent symptoms were more likely to be post-menopausal. Women who reported persistent symptoms or met criteria for remitted PTSD had slightly higher BMI and were more likely to have a history of hypertension.

**Table 1.** Participant characteristics by PTSD and trauma status (total sample N=2950), assessed at or near time of blood collection.

|  | Full Sample<br>n=2950 | With<br>persistent<br>PTSD<br>symptoms<sup>1</sup><br>n=817 | With remitted<br>probable<br>lifetime PTSD<br>n=235 | Trauma<br>exposed<br>without<br>PTSD<br>n=1170 | Trauma<br>unexposed<br>n=728 | Missing |
| --- | --- | --- | --- | --- | --- | --- |
| ***Demographic and technical factors*** |  |  |  |  |  |  |
| Age at blood collection, year, M(SD) | 47.3 (7.0) | 47.7 (7.1) | 47.1 (7.0) | 47.4 (7.2) | 46.7 (6.7) | 0 |
| Race/ethnicity: non-White, N(%) | 67 (2.3) | 17 (2.1) | 2 (0.9) | 30 (2.6) | 18 (2.5) | 0 |
| Menopausal status, N(%) |  |  |  |  |  | 0 |
| Pre-menopausal | 1810 (61.4) | 465 (56.9) | 145 (61.7) | 727 (62.1) | 473 (65.0) |  |
| Post-menopausal | 802 (27.2) | 249 (30.5) | 60 (25.5) | 306 (26.2) | 187 (25.7) |  |
| Missing/Unknown | 338 (11.5) | 103 (12.6) | 30 (12.8) | 137 (11.7) | 68 (9.3) |  |
| Fasting status: non-fasting, N(%) | 722 (24.5) | 194 (23.7) | 56 (23.8) | 290 (24.8) | 182 (25.0) | 0 |
| ***Medical factors*** |  |  |  |  |  |  |
| Statin or other lipid lowering drug use, N(%) | 331 (11.2) | 111 (13.6) | 29 (12.3) | 119 (10.2) | 72 (9.9) | 0 |
| Hormone therapy current use, N(%) | 419 (14.5) | 132 (16.4) | 40 (17.3) | 148 (13.0) | 99 (13.7) | 1.9 |
| Hypertension history, N(%) | 428 (14.5) | 138 (16.9) | 41 (17.4) | 151 (12.9) | 98 (13.5) | 0 |
| Type 2 diabetes history, N(%) | 55 (1.9) | 21 (2.6) | 6 (2.6) | 15 (1.3) | 13 (1.8) | 0 |
| ***Biobehavioral factors*** |  |  |  |  |  |  |
| Diet quality score<sup>2</sup>, M(SD) | 50.5 (12.6) | 51.3 (12.9) | 50.2 (12.6) | 50.5 (13.0) | 49.7 (11.6) | 0.2 |
| Recreational physical activity in MET-hrs/wk<sup>3</sup>,<br>M(SD) | 19.3 (21.8) | 18.2 (20.7) | 21.4 (30.3) | 19.9 (21.7) | 18.9 (19.6) | 0.8 |
| Alcohol intake, g/day, M(SD) | 4.3 (7.7) | 4.0 (7.4) | 3.9 (6.1) | 4.4 (8.3) | 4.5 (7.5) | 4 |
| Current smoker, N(%) | 201 (6.8) | 66 (8.1) | 21 (8.9) | 73 (6.2) | 41 (5.6) | 0 |
| Caffeine intake, mg/day, M(SD) | 218.9 (207.1) | 222.8 (215.7) | 217.3 (209.3) | 222.9 (212.2) | 208.5 (187.3) | 0.5 |
| Body mass index, M(SD) | 25.9 (5.7) | 26.5 (6.1) | 26.5 (5.9) | 25.8 (5.6) | 25.1 (5.0) | 0 |

<sup>1</sup> Women with any persistent PTSD symptoms are grouped together for the purpose of showing the covariate distribution in this table, but the continuous symptom count was included in modeling.
<sup>2</sup> Measured by the Alternative Healthy Eating Index (AHEI).
<sup>3</sup> Total metabolic equivalent task (MET) hours per week.

### Does PTSD share common metabolic alterations with depression and anxiety?

A minimally adjusted meta-analysis of the association between the MDS and PTSD/trauma exposure demonstrated a significant positive relationship between persistent PTSD symptoms and the MDS; for each additional persistent symptom, MDS increased by 0.08 SD on average (95% CI: [0.04,0.13]) (**Table 2**). However, MDS was not associated with remitted PTSD (β=0.14, [-0.12, 0.39]) or trauma (β=-0.06, [-0.38, 0.27]). The association with persistent symptoms remained significant after adjusting for medical and biobehavioral factors in Model 3 (β=0.07, [0.02, 0.11]).

**Table 2.** Associations between persistent PTSD symptoms and the metabolite-based distress score.

| Exposure | Model 1: Minimal |  | Model 2: Medical |  | Model 3: Biobehavioral |  |
| --- | --- | --- | --- | --- | --- | --- |
| | $\beta$ (95% CI) | P-value | $\beta$ (95% CI) | P-value | $\beta$ (95% CI) | P-value |
| ***Random-Effects Meta-Analysis (N=2950)*** |  |  |  |  |  |  |
| Persistent PTSD Symptom Count | **0.08 (0.04, 0.13)** | **5.00E-04** | **0.08 (0.03, 0.12)** | **0.0011** | **0.07 (0.02, 0.11)** | **0.0036** |
| Remitted PTSD | 0.14 (-0.12, 0.39) | 0.2927 | 0.12 (-0.13, 0.37) | 0.3367 | 0.1 (-0.14, 0.34) | 0.4178 |
| Trauma Exposure | -0.06 (-0.38, 0.27) | 0.7212 | -0.03 (-0.32, 0.27) | 0.8616 | -0.1 (-0.42, 0.22) | 0.5356 |
| ***NHSII Merged Dataset (N=2550)*** |  |  |  |  |  |  |
| Persistent PTSD Symptom Count | **0.06 (0.01, 0.12)** | **0.0252** | **0.06 (0.01, 0.12)** | **0.0313** | 0.05 (-0.01, 0.1) | 0.0831 |
| Remitted PTSD | 0.11 (-0.17, 0.39) | 0.4494 | 0.12 (-0.16, 0.4) | 0.4146 | 0.05 (-0.23, 0.32) | 0.7466 |
| Trauma Exposure | 0.13 (-0.02, 0.28) | 0.0969 | 0.14 (-0.02, 0.29) | 0.078 | 0.09 (-0.06, 0.23) | 0.258 |
| ***Mind-Body Study (N=204)*** |  |  |  |  |  |  |
| Persistent PTSD Symptom Count | **0.18 (0.03, 0.33)** | **0.018** | **0.17 (0.03, 0.31)** | **0.0196** | **0.16 (0.02, 0.29)** | **0.0265** |
| Remitted PTSD | 0.77 (-0.19, 1.72) | 0.1164 | 0.64 (-0.27, 1.55) | 0.1679 | 0.7 (-0.17, 1.57) | 0.1144 |
| Trauma Exposure | -0.23 (-0.82, 0.36) | 0.441 | -0.24 (-0.8, 0.32) | 0.405 | -0.18 (-0.73, 0.36) | 0.5122 |
| ***Severe Distress Sample (N=196)*** |  |  |  |  |  |  |
| Persistent PTSD Symptom Count | 0.1 (-0.01, 0.2) | 0.0662 | 0.08 (-0.02, 0.18) | 0.1231 | 0.08 (-0.02, 0.17) | 0.11 |
| Remitted PTSD | -0.03 (-0.68, 0.62) | 0.9345 | -0.11 (-0.74, 0.52) | 0.7321 | 0.07 (-0.53, 0.67) | 0.8237 |
| Trauma Exposure | -0.33 (-0.84, 0.18) | 0.2008 | -0.25 (-0.74, 0.24) | 0.3261 | -0.41 (-0.88, 0.06) | 0.0893 |

Persistent PTSD symptoms, lifetime PTSD in remission, and lifetime trauma exposure are included in the models simultaneously.
Bold face indicates statistical significance ( $p < 0.05$ ).

Four individual metabolites comprising the MDS were significantly associated (FDR<0.05, correcting for 19 tests) with higher persistent PTSD symptoms in minimally adjusted models, and three remained significant after accounting for medical factors (Model 2 in **Figure 1** and **Supplemental Table S1**): lower serotonin (β=-0.04 [−0.07, −0.01]) and higher levels of two glycerophospholipid metabolites (C18:0 LPE: β=0.04 [0.02, 0.07]; C34:3 PC: β=0.08 [0.02, 0.13]). The directions of these associations were consistent with previously reported associations with depression or anxiety. No associations were found with remitted PTSD or trauma exposure (**Supplemental Tables S1B&C)**.

**Figure 1.**
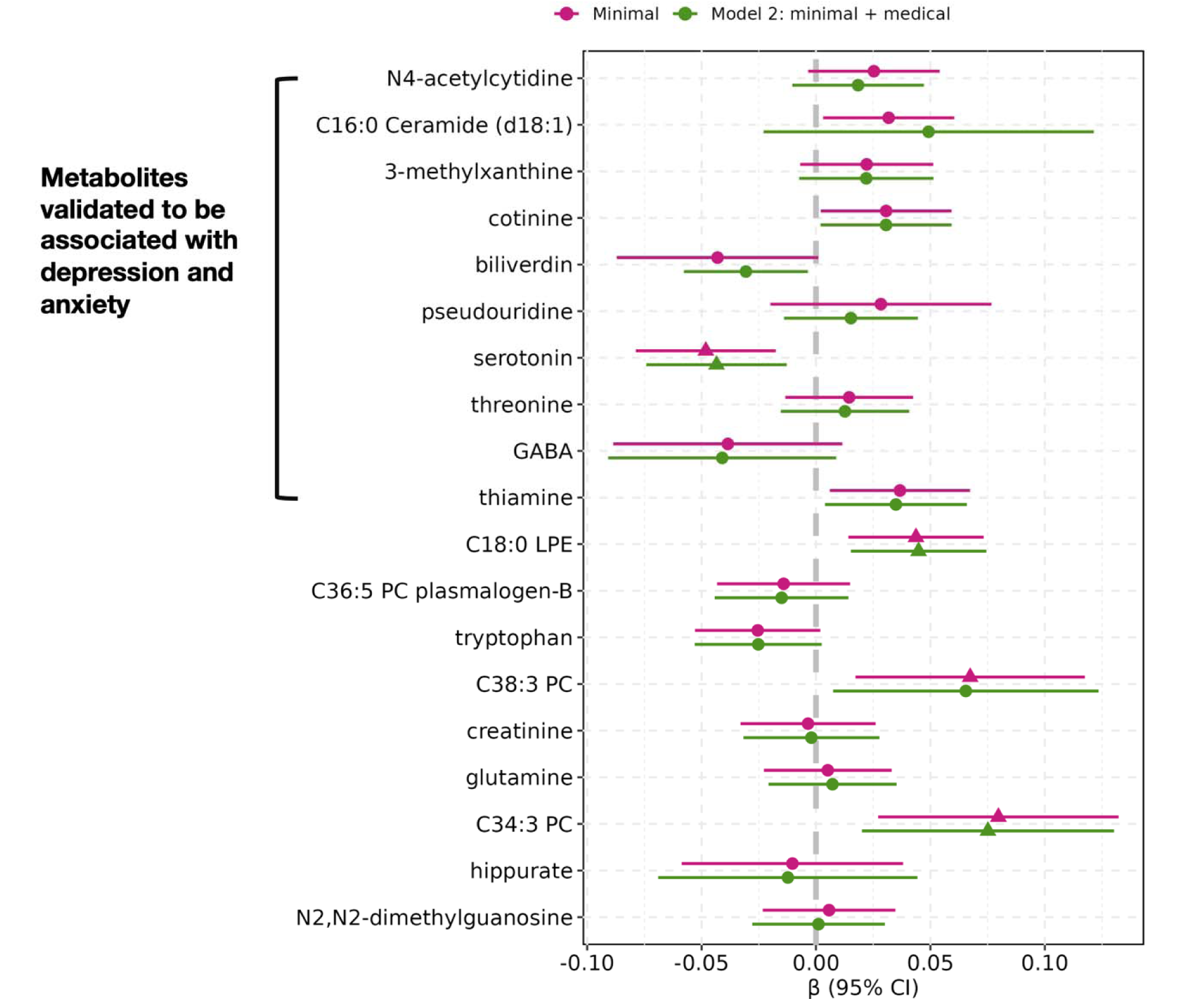
Associations between persistent PTSD symptoms and 19 metabolites previously associated with depression and anxiety included in the metabolite-based distress score (MDS), adjusting for baseline covariates (minimal, or Model 1) and medical factors (Model 2). *Note.* Estimates are obtained from random effects meta-analyses of all three subsamples. For the β estimates, triangles indicate statistically significant associations after false discovery rate corrections for the number of metabolites tested (19; FDR adjusted p<0.05) and circles indicate associations not statistically significant. Statistical model: metabolite-based distress score ∼ Persistent PTSD symptom count + remitted PTSD + trauma exposure + covariates Model 1 (minimal) covariates: age, race/ethnicity, menopausal status, and fasting status at blood collection. Model 2 covariates: Model 1 + use of statins or other lipid lowering drugs, hormone therapy, hypertension history, and type 2 diabetes history.

In the metabolite set enrichment analysis of metabolite classes based on metabolome-wide results, glycolipids and glycerophospholipids were significantly enriched for positive associations with persistent PTSD symptoms (**Supplemental Figure S2**). However, glycerophospholipids were enriched for inverse associations with depression/anxiety.

Sphingolipids, imidazopyrimidines, and steroids were significantly enriched for associations with depression/anxiety; while the direction of enrichment for these three classes were the same with persistent PTSD symptoms, they did not reach significance after FDR corrections.

### Are unique metabolic markers related to PTSD symptoms?

In agnostic analyses evaluating 339 metabolites (including metabolites in the MDS), 29 were significantly linked to persistent PTSD symptoms in the minimally adjusted (Model 1) meta-analyses after multiple testing corrections (FDR<0.05), and no metabolites were linked to trauma exposure or remitted PTSD (**Supplemental Table S2**).

Nine metabolites remained significantly associated with persistent PTSD symptoms after accounting for medical factors (Model 2 in **Figure 2** and **Supplemental Table S2**). None of these metabolites showed significant between-sample heterogeneity (p>0.26 for Cochran’s Q), despite age-associated differences in demographic and health characteristics between subsamples. Eight metabolites were positively associated with persistent symptoms, and one was inversely associated (C5:1 carnitine). There was no overlap between these nine metabolites and the previously identified MDS metabolites. The magnitude of coefficients for each additional persistent symptom ranged from 0.05 (95% CI [0.02, 0.08], C36:1 PE) to 0.08 (95% CI [0.04, 0.12], homoarginine). The most common metabolite class represented among these nine metabolites was glycerophospholipids (7 out of 9). Among these nine metabolites identified in Model 2, all but homoarginine remained significant after adjusting further for biobehavioral factors (Model 3).

**Figure 2.**
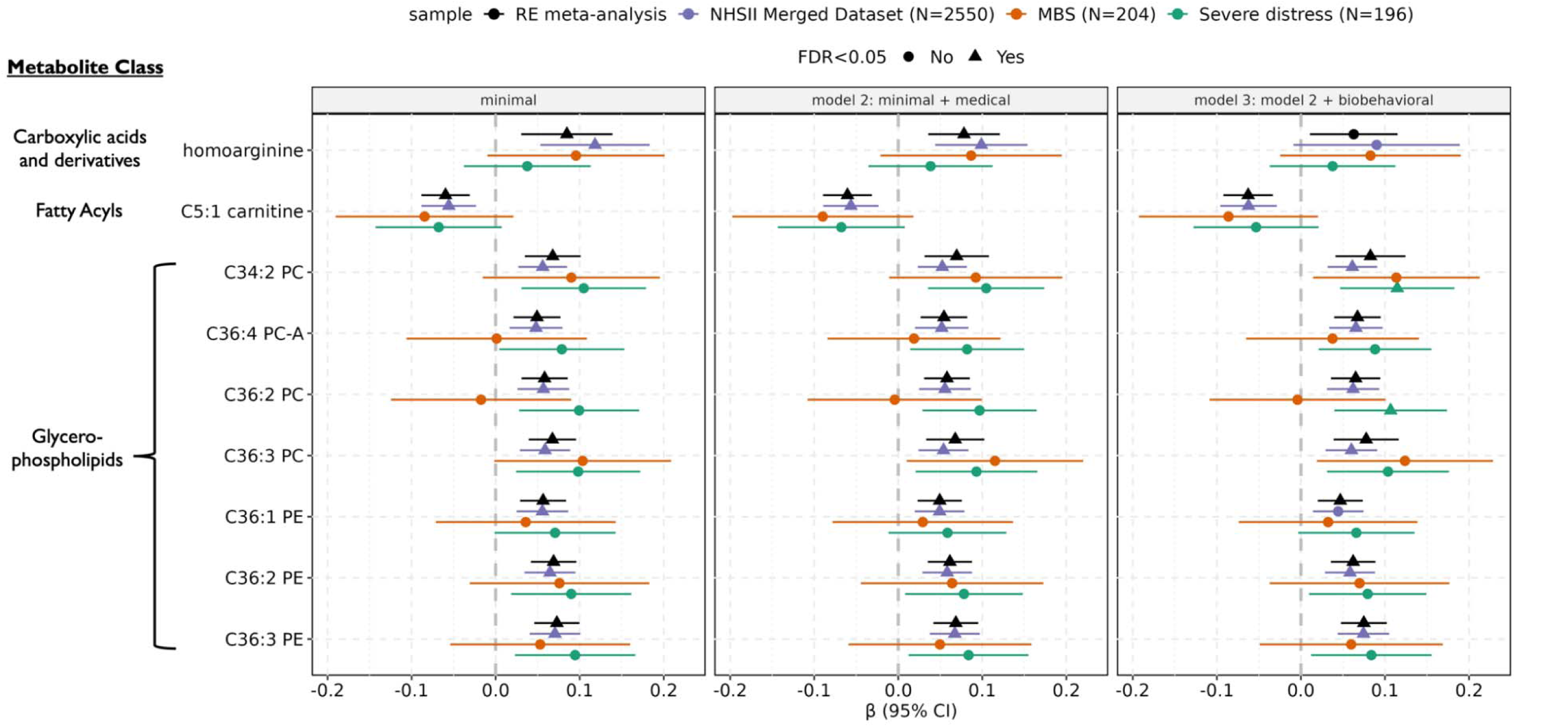
Agnostic per metabolite analysis results: statistically significant associations between persistent PTSD symptoms and nine metabolites in the random-effects meta-analysis, after adjusting for technical and medical covariates (Model 2) and multiple testing corrections. *Note.* Estimates are obtained from both random effects meta-analyses of all three subsamples (black), and individual sub-samples: NHSII merged dataset (purple), the Mind-Body Study (MBS; orange), and the Severe Distress Sample (green). For the β estimates, triangles indicate statistically significant associations after false discovery rate corrections for the number of metabolites tested (339; FDR adjusted p<0.05) and circles indicate associations not statistically significant. Statistical model: metabolite ∼ Persistent PTSD symptom count + remitted PTSD + trauma exposure + covariates Model 1 (minimal) covariates: age, race/ethnicity, menopausal status, fasting status at blood collection, and sub-study indicator (only applicable in the NHSII Merged Dataset); Model 2 covariates: Model 1 + use of statins or other lipid lowering drugs, hormone therapy, hypertension history, and type 2 diabetes history; Model 3 covariates: Model 2 + diet quality, physical activity, alcohol intake, smoking status, caffeine intake, and body-mass index.

### Differential Network Analysis

Informed by the agnostic analysis where associations were evident only with respect to persistent PTSD symptoms in all models, we estimated a differential network comparing participants with (N=272) versus without (N=586) any persistent PTSD symptoms (**Figure 3**). Out of 29 metabolites included in the analysis, 17 with 54 significant edges (FDR<0.05) between them were retained in the graph. Half of the significant edges were positive, suggesting the partial correlation between metabolite pairs was higher among participants with versus without persistent symptoms, with the largest difference observed between C50:4 TAG and C34:2 PC (Δ_partial r_=0.16). Among the negative edges (i.e., edges representing lower partial correlations among participants with persistent symptoms), the largest difference was observed between C22:0 Ceramide (d18:1) and C50:4 TAG (Δ_partial r_=-0.09). Summary results of network measures highlight the importance of a triglyceride, C50:4 TAG, in the network of metabolite differential partial correlations, which had the highest hub score, betweenness, and closeness **(Supplemental Table S3**).

**Figure 3.**
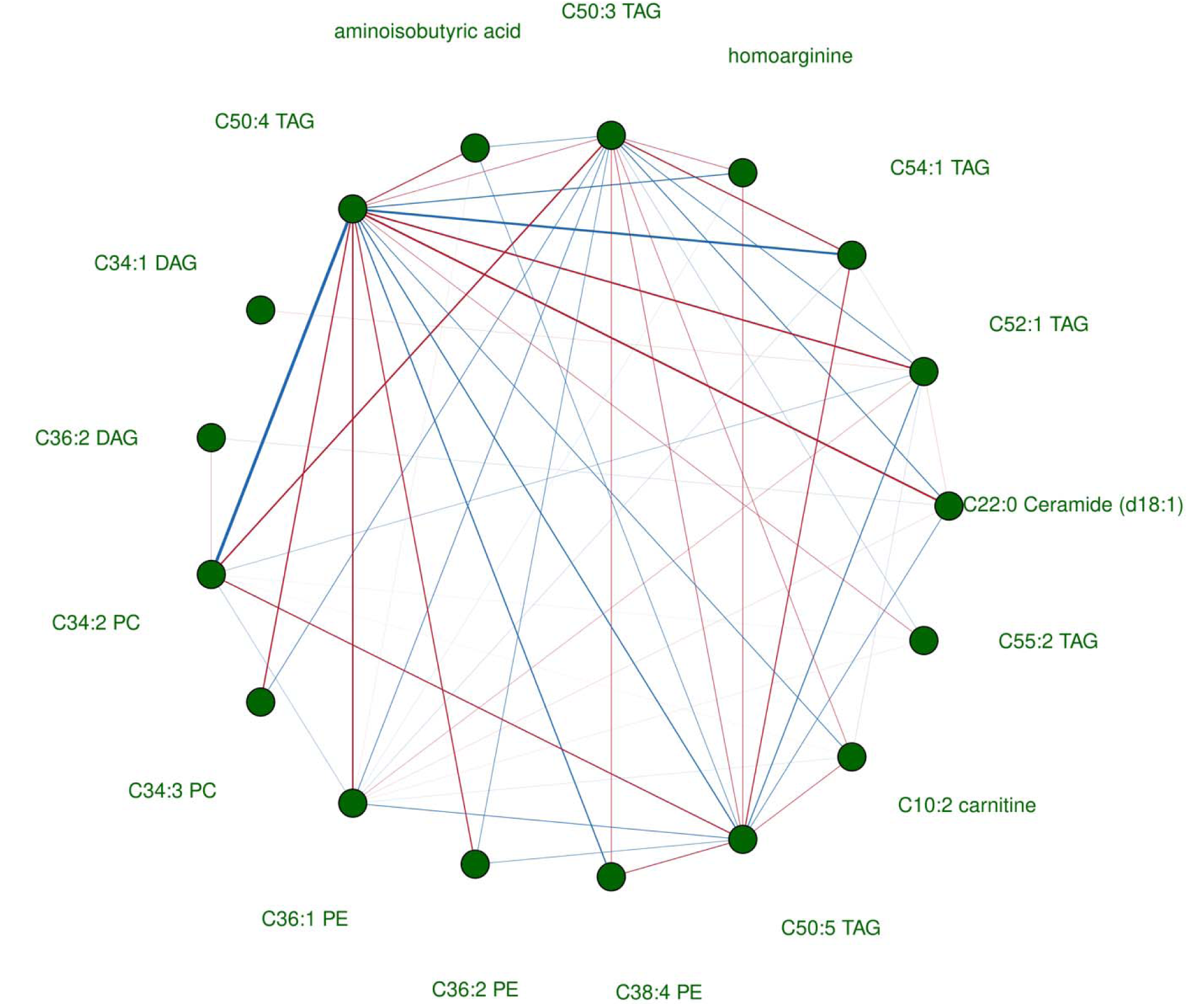
Differential network showing statistically significant differences in partial correlations estimated between 29 metabolites associated with persistent PTSD symptoms in the meta-analysis of the minimal model. Seventeen metabolites connected to 54 significant edges after multiple testing corrections (FDR<0.05) are shown in the figure. *Note.* Analysis was performed within a subsample of 858 independent participants who had complete data on all 29 metabolites selected based on the minimally adjusted agnostic analysis. The differential network was constructed by comparing participants reporting persistent PTSD symptoms in 2008 (N=272) and participants who did not report persistent symptoms regardless of trauma exposure status (N=586). Red edges indicate that partial correlations between pairs of metabolites were higher among participants with persistent symptoms versus without. Blue edges indicate that partial correlations between pairs of metabolites were lower among participants with persistent symptoms versus without. The width of edges corresponds to the weight, i.e., larger difference in partial correlations between individuals with persistent PTSD symptoms versus without.

## Discussion

Our study is the first large-scale metabolomics analysis of PTSD in a predominantly civilian sample of middle-aged women. We found overlap as well as notable differences in metabolic alterations linked to PTSD in comparison to metabolites previously linked with depression or anxiety in this population. We identified nine metabolites associated with persistent PTSD symptoms but not linked to depression and anxiety in prior research. Considering patterns of correlations in a metabolite network, triglycerides may be key markers in metabolomic differences associated with persistent PTSD.

Consistent with prior investigations of metabolic alterations and distress, the current study highlights the importance of phospholipid dysregulation in relation to persistent PTSD. Phospholipids form the backbone of neural membranes and play a central role in maintaining membrane functions and transmembrane signaling in the brain (Farooqui et al., 2004). A recent study of PTSD in mice identified sustained increased hippocampal levels of glycerophosphoethanolamines (PEs) and phosphatidylcholine (PCs) in mice characterized as being susceptible to PTSD (Pascual Cuadrado et al., 2022). Prior studies have also linked phospholipid dysregulation reflected in human peripheral tissues (e.g., blood) to higher risks of neurodegenerative diseases (González-Domínguez et al., 2014; Li et al., 2019) and inflammation (Leitinger, 2003). Although several studies have identified differences in phospholipid concentrations related to PTSD, the direction of effects varies: similar to our findings, Karabatsiakis et al. found elevated levels of four PEs among a small sample of PTSD patients (N=38) in clinical, civilian settings (Karabatsiakis et al., 2015). A metabolomic analysis in two samples of male veterans (combined N=204) also pointed to increased levels of phospholipids across several subclasses (Konjevod et al., 2021). However, two studies among male veterans (N=476) (Konjevod et al., 2022) and male US active duty soldiers (N=120) (Emmerich et al., 2016) revealed *decreased* levels of PEs and PCs in PTSD cases compared to healthy controls. Intriguingly, a recent lipidomic analysis in 80 men and women reported only one lipid subclass, PEs, was altered among women with PTSD whereas 9 out of 13 subclasses were altered among men with PTSD (Bhargava et al., 2024). These differences may be due to variations in trauma type, study population, and analytic strategies; with data from a large, civilian sample, our study may better capture subclinical variations in PTSD symptom severity and nuanced differences in associated metabolomics profiles. Future longitudinal investigations in well-characterized, large samples including men and women may help characterize lipid dysregulations following the development of PTSD more precisely and identify key effect modifiers, such as gender differences in trauma responses (Olff, 2017).

Another novel contribution of our study is identifying a positive association with homoarginine and an inverse association with C5:1 carnitine in relation to persistent PTSD symptoms. Homoarginine may promote synthesis of nitric oxide and increase oxidative stress (Bernstein et al., 2015; Lambert et al., 1992). The positive association with homoarginine may reflect an increase in nitric oxide formation and inflammation occurring with PTSD (Oosthuizen et al., 2005). Some carnitines, namely, pentadecanoylcarnitine, have also shown endocannabinoid-like activities and been implicated in regulating inflammation and mood (Venn-Watson et al., 2022). Interestingly, in a pilot study of chronically stressed female mice, carnitines were over-expressed in plasma but under-expressed in ovarian tissues, highlighting the dynamic processes of uptake or release of carnitine metabolites under stress in different tissues (Zeleznik et al., 2022). Our findings suggest that homoarginine and C5:1 carnitine may be promising targets for follow-up experiments of stress response pathophysiology.

The substantial evidence for metabolite associations with persistent PTSD symptoms but not trauma exposure or remitted PTSD suggests chronic, non-remitted symptoms may have more profound metabolic consequences. The same pattern has been observed in PTSD research examining other biomarkers: past studies have found elevated inflammation among participants with current PTSD but fewer differences between individuals with remitted PTSD versus those who never experienced PTSD (Glaus et al., 2018; O’Donovan et al., 2017). However, findings with respect to associations with trauma or remitted PTSD in our study should be interpreted with the comparison groups in mind. Per our modeling strategies, women who were trauma-exposed but reported no lifetime PTSD were compared to women who reported no trauma exposure. Prior work examining any trauma exposure more broadly (regardless of PTSD symptoms) has identified metabolite associations with trauma and pointed to the mediating roles of psychological distress (Huang et al., 2022).

Our comparison of PTSD-related metabolic alterations with other highly comorbid chronic distress phenotypes, namely, depression or anxiety, suggests some alterations in metabolic markers may be specific to each disorder. Substantial overlap exists between known underlying biological changes occurring with PTSD and depression, including structural and functional changes in the brain and endocrine responses (Ploski & Vaidya, 2021). Yet there are notable distinctions as well, such as enhanced glucocorticoid sensitivity in PTSD versus greater glucocorticoid resistance in major depression (Yehuda, 2001). Our findings shed light on shared metabolic features across disorders, such as decreased serotonin and relevance of phospholipid metabolism (Lin et al., 2010). We also highlight the possibility of disorder-specific markers, such as elevated levels of glycerolipid and glycerophospholipid metabolites in PTSD, and dysregulations of amino acids and fatty acids occurring with depression and anxiety (Shutta et al., 2021). Understanding whether these distinctions between disorders replicate across populations and identifying transdiagnostic metabolomic profiles of distress are critical next steps (Pinto et al., 2017; Waszczuk et al., 2023), especially considering the challenges and failures to validate candidate biomarkers for several major psychiatric disorders (Abi-Dargham et al., 2023).

These findings may further provide insight into mechanisms underlying observed elevated cardiometabolic disease risk associated with PTSD. as well as mortality (Levine et al., 2013; Roberts et al., 2020). In a recent analysis of plasma metabolites associated with mortality and longevity among over 11,000 US adults, three (C36:3 PC, C36:2 PE, and C36:3 PE) out of the nine metabolites linked to PTSD symptoms in our study also showed positive associations with higher all-cause mortality risk (Wang et al., 2023). Immune alterations in response to traumatic stress and downstream behavioral changes may contribute to lipid accumulation (Mellon et al., 2018). Dysregulation of phospholipid metabolism can give rise to mitochondrial dysfunction and consequently heightened oxidative stress, which in turn could contribute to impaired insulin signaling, perpetuating a vicious cycle that leads to elevated risk of cardiometabolic diseases (Meikle & Summers, 2017). While diet and pharmacological agents have been the most studied intervention targets, the prominent lipid associations with PTSD suggest that preventing, screening, and treating psychological distress could be another strategy for improving cardiometabolic health (Levine et al., 2021).

Our study has several notable strengths. First, most existing metabolomic studies of PTSD were based on male, military, and/or highly selected clinical samples (Zhu et al., 2022); our findings address an important gap by examining a large, population-based sample of predominantly civilian women. Second, we strengthened the methodological rigor of our study by leveraging detailed covariate data collected over decades and combining measures from blood samples collected at different time points relative to PTSD assessments. Nonetheless, several limitations should be considered. First, the NHSII cohort consists of predominantly White, professional women; as such, our findings may not generalize to populations experiencing more severe forms of traumatic events or PTSD symptoms and women with non-White racial and ethnic backgrounds. Relatedly, although we did not observe any age-dependent between-sample heterogeneity according to the Cochran’s Q-tests, these tests may be underpowered when the number of substudies is small (Gavaghan et al., 2000). Therefore, we cannot rule out potential effect modification by age or other demographic characteristics. Third, trauma and PTSD measures were assessed once in 2008 whereas blood collection occurred between 1996 and 1999 for most samples, which could introduce recall bias. For women whose blood samples were collected after 2008, exposure misclassification was also a concern because no PTSD information was available after 2008. To mitigate these potential biases, we included samples with later blood collections when possible, carefully dated PTSD onset by checking the reported age of trauma exposure, and interpreted our findings cautiously. Nonetheless, the identified associations may reflect reverse causation whereby changes in metabolite levels increased PTSD susceptibility. Moreover, we cannot fully disentangle effects of PTSD and depression on metabolomic profiles: depression and PTSD are highly comorbid and identifying their separate effects would require being able to specify a temporal causal relationship between the development of the conditions and test this causal model using repeated assessments of PTSD and depression, which were unavailable in our study. Given the constraints, we focused on comparing the metabolites associated with PTSD versus metabolites linked to depression in previous publications using independent samples.

Through large-scale plasma metabolomic profiling of PTSD among middle-aged women, we identified several promising metabolic markers associated with having persistent PTSD symptoms. Metabolic dysregulations, in particular elevated lipid levels, may occur with PTSD and be partly responsible for associations of PTSD with adverse physical health outcomes. With further research, screening for and treating chronic distress disorders like PTSD could be a crucial intervention for improving overall health among trauma-exposed women.

## Data Availability

All data produced in the present study are available upon reasonable request to the authors.

## Acknowledgement

We thank Dr. Shaili C. Jha (Department of Epidemiology, Harvard T.H. Chan School of Public Health) for her contribution to data collection and management. We thank Drs. Karestan C. Koenen, Liming Liang, Lori B. Chibnik (Department of Epidemiology, Harvard T.H. Chan School of Public Health), and Karmel W. Choi (Department of Psychiatry, Massachusetts General Hospital) for their insightful feedback on study results. We also thank Rebekah Kristal (Department of Biostatistics and Epidemiology, University of Massachusetts Amherst) for her feedback on the manuscript. We thank the participants and the staff of the Nurses’ Health Study II (NHSII) for their valuable contributions.

## References

Abi-Dargham, A., Moeller, S. J., Ali, F., DeLorenzo, C., Domschke, K., Horga, G., Jutla, A., Kotov, R., Paulus, M. P., Rubio, J. M., Sanacora, G., Veenstra-VanderWeele, J., & Krystal, J. H. (2023). Candidate biomarkers in psychiatric disorders: State of the field. World Psychiatry, 22(2), 236–262. 10.1002/wps.21078

Ainsworth, B. E., Haskell, W. L., Leon, A. S., Jacobs, D. R., Montoye, H. J., Sallis, J. F., & Paffenbarger, R. S. (1993). Compendium of physical activities: Classification of energy costs of human physical activities. Medicine and Science in Sports and Exercise, 25(1), 71–80. 10.1249/00005768-199301000-00011

American Psychiatric Association. (1994). Diagnostic criteria from DSM-IV. The Association.

Balasubramanian, R., Shutta, K. H., Guasch-Ferre, M., Huang, T., Jha, S. C., Zhu, Y., Shadyab, A. H., Manson, J. E., Corella, D., Fitó, M., Hu, F. B., Rexrode, K. M., Clish, C. B., Hankinson, S. E., & Kubzansky, L. D. (2023). Metabolomic profiles of chronic distress are associated with cardiovascular disease risk and inflammation-related risk factors. Brain, Behavior, and Immunity. 10.1016/j.bbi.2023.08.010

Bernstein, H.-G., Jäger, K., Dobrowolny, H., Steiner, J., Keilhoff, G., Bogerts, B., & Laube, G. (2015). Possible sources and functions of l-homoarginine in the brain: Review of the literature and own findings. Amino Acids, 47(9), 1729–1740. 10.1007/s00726-015-1960-y

Bhargava, A., Knapp, J. D., Fiehn, O., Neylan, T. C., & Inslicht, S. S. (2024). An exploratory study on lipidomic profiles in a cohort of individuals with posttraumatic stress disorder. Scientific Reports, 14(1), 15256. 10.1038/s41598-024-62971-7

Bot, M., Milaneschi, Y., Al-Shehri, T., Amin, N., Garmaeva, S., Onderwater, G. L. J., Pool, R., Thesing, C. S., Vijfhuizen, L. S., Vogelzangs, N., Arts, I. C. W., Demirkan, A., Duijn, C. van, Greevenbroek, M. van, Kallen, C. J. H. van der, Köhler, S., Ligthart, L., Maagdenberg, A. M. J. M. van den, Mook-Kanamori, D. O.,…Sattar, N. (2020). Metabolomics Profile in Depression: A Pooled Analysis of 230 Metabolic Markers in 5283 Cases With Depression and 10,145 Controls. Biological Psychiatry, 87(5), 409–418. 10.1016/j.biopsych.2019.08.016

Boulet, M. M., Chevrier, G., Grenier-Larouche, T., Pelletier, M., Nadeau, M., Scarpa, J., Prehn, C., Marette, A., Adamski, J., & Tchernof, A. (2015). Alterations of plasma metabolite profiles related to adipose tissue distribution and cardiometabolic risk. American Journal of Physiology-Endocrinology and Metabolism, 309(8), E736–E746. 10.1152/ajpendo.00231.2015

Brandes, U. (2001). A faster algorithm for betweenness centrality*. The Journal of Mathematical Sociology, 25(2), 163–177. 10.1080/0022250X.2001.9990249

Breslau, N., Peterson, E. L., Kessler, R. C., & Schultz, L. R. (1999). Short Screening Scale for DSM-IV Posttraumatic Stress Disorder. American Journal of Psychiatry, 156(6), 908– 911. 10.1176/ajp.156.6.908

Chiuve, S. E., Fung, T. T., Rimm, E. B., Hu, F. B., McCullough, M. L., Wang, M., Stampfer, M. J., & Willett, W. C. (2012). Alternative Dietary Indices Both Strongly Predict Risk of Chronic Disease,. The Journal of Nutrition, 142(6), 1009–1018. 10.3945/jn.111.157222

Clish, C. B. (2015). Metabolomics: An emerging but powerful tool for precision medicine. Cold Spring Harbor Molecular Case Studies, 1(1), a000588. 10.1101/mcs.a000588

Davyson, E., Shen, X., Gadd, D. A., Bernabeu, E., Hillary, R. F., McCartney, D. L., Adams, M., Marioni, R., & McIntosh, A. M. (2023). Metabolomic investigation of major depressive disorder identifies a potentially causal association with polyunsaturated fatty acids. Biological Psychiatry. 10.1016/j.biopsych.2023.01.027

Do, K. T., Wahl, S., Raffler, J., Molnos, S., Laimighofer, M., Adamski, J., Suhre, K., Strauch, K., Peters, A., Gieger, C., Langenberg, C., Stewart, I. D., Theis, F. J., Grallert, H., Kastenmüller, G., & Krumsiek, J. (2018). Characterization of missing values in untargeted MS-based metabolomics data and evaluation of missing data handling strategies. Metabolomics, 14(10), 128. 10.1007/s11306-018-1420-2

Dunn, W. B., Bailey, N. J. C., & Johnson, H. E. (2005). Measuring the metabolome: Current analytical technologies. The Analyst, 130(5), 606. 10.1039/b418288j

Emmerich, T., Abdullah, L., Crynen, G., Dretsch, M., Evans, J., Ait-Ghezala, G., Reed, J., Montague, H., Chaytow, H., Mathura, V., Martin, J., Pelot, R., Ferguson, S., Bishop, A., Phillips, J., Mullan, M., & Crawford, F. (2016). Plasma Lipidomic Profiling in a Military Population of Mild Traumatic Brain Injury and Post-Traumatic Stress Disorder with Apolipoprotein E 4–Dependent Effect. Journal of Neurotrauma, 33(14), 1331–1348. 10.1089/neu.2015.4061

Farooqui, A. A., Ong, W.-Y., & Horrocks, L. A. (2004). Biochemical Aspects of Neurodegeneration in Human Brain: Involvement of Neural Membrane Phospholipids and Phospholipases A2. Neurochemical Research, 29(11), 1961–1977. 10.1007/s11064-004-6871-3

Freeman, L. C. (1978). Centrality in social networks conceptual clarification. Social Networks, 1(3), 215–239. 10.1016/0378-8733(78)90021-7

Fujisaka, S., Avila-Pacheco, J., Soto, M., Kostic, A., Dreyfuss, J. M., Pan, H., Ussar, S., Altindis, E., Li, N., Bry, L., Clish, C. B., & Kahn, C. R. (2018). Diet, Genetics, and the Gut Microbiome Drive Dynamic Changes in Plasma Metabolites. Cell Reports, 22(11), 3072– 3086. 10.1016/j.celrep.2018.02.060

Galatzer-Levy, I. R., Nickerson, A., Litz, B. T., & Marmar, C. R. (2013). Patterns of Lifetime Ptsd Comorbidity: A Latent Class Analysis. Depression and Anxiety, 30(5), 489–496. 10.1002/da.22048

Gavaghan, D. J., Moore, R. A., & McQuay, H. J. (2000). An evaluation of homogeneity tests in meta-analyses in pain using simulations of individual patient data. Pain, 85(3), 415–424. 10.1016/S0304-3959(99)00302-4

Glaus, J., von Känel, R., Lasserre, A. M., Strippoli, M.-P. F., Vandeleur, C. L., Castelao, E., Gholam-Rezaee, M., Marangoni, C., Wagner, E.-Y. N., Marques-Vidal, P., Waeber, G., Vollenweider, P., Preisig, M., & Merikangas, K. R. (2018). The bidirectional relationship between anxiety disorders and circulating levels of inflammatory markers: Results from a large longitudinal population-based study. Depression and Anxiety, 35(4), 360–371. 10.1002/da.22710

González-Domínguez, R., García-Barrera, T., & Gómez-Ariza, J. L. (2014). Combination of metabolomic and phospholipid-profiling approaches for the study of Alzheimer’s disease. Journal of Proteomics, 104, 37–47. 10.1016/j.jprot.2014.01.014

Ha, M. J., Baladandayuthapani, V., & Do, K.-A. (2015). DINGO: Differential network analysis in genomics. Bioinformatics, 31(21), 3413–3420. 10.1093/bioinformatics/btv406

Huang, T., Balasubramanian, R., Yao, Y., Clish, C. B., Shadyab, A. H., Liu, B., Tworoger, S. S., Rexrode, K. M., Manson, J. E., Kubzansky, L. D., & Hankinson, S. E. (2020). Associations of depression status with plasma levels of candidate lipid and amino acid metabolites: A meta-analysis of individual data from three independent samples of US postmenopausal women. Molecular Psychiatry. 10.1038/s41380-020-00870-9

Huang, T., Trudel-Fitzgerald, C., Poole, E. M., Sawyer, S., Kubzansky, L. D., Hankinson, S. E., Okereke, O. I., & Tworoger, S. S. (2019). The Mind–Body Study: Study design and reproducibility and interrelationships of psychosocial factors in the Nurses’ Health Study II. Cancer Causes & Control, 30(7), 779–790. 10.1007/s10552-019-01176-0

Huang, T., Zeleznik, O. A., Roberts, A. L., Balasubramanian, R., Clish, C. B., Eliassen, A. H., Rexrode, K. M., Tworoger, S. S., Hankinson, S. E., Koenen, K. C., & Kubzansky, L. D. (2022). Plasma Metabolomic Signature of Early Abuse in Middle-Aged Women. Psychosomatic Medicine, 84(5), 536–546. 10.1097/PSY.0000000000001088

Huang, T., Zhu, Y., Shutta, K. H., Balasubramanian, R., Zeleznik, O. A., Rexrode, K. M., Clish, C. B., Sun, Q., Hu, F. B., Kubzansky, L. D., & Hankinson, S. E. (2023). A Plasma Metabolite Score Related to Psychological Distress and Diabetes Risk: A Nested Case-control Study in US Women. The Journal of Clinical Endocrinology & Metabolism, dgad731. 10.1210/clinem/dgad731

Karabatsiakis, A., Hamuni, G., Wilker, S., Kolassa, S., Renu, D., Kadereit, S., Schauer, M., Hennessy, T., & Kolassa, I.-T. (2015). Metabolite profiling in posttraumatic stress disorder. Journal of Molecular Psychiatry, 3(1), 2. 10.1186/s40303-015-0007-3

Kilpatrick D. G., Resnick H. S., Milanak M. E., Miller M. W., Keyes K. M., & Friedman M. J. (2013). National Estimates of Exposure to Traumatic Events and PTSD Prevalence Using DSM-IV and DSM-5 Criteria. Journal of Traumatic Stress, 26(5), 537–547. 10.1002/jts.21848

Kleinberg, J. M. (1999). Authoritative sources in a hyperlinked environment. Journal of the ACM, 46(5), 604–632. 10.1145/324133.324140

Konjevod, M., Nedic Erjavec, G., Nikolac Perkovic, M., Sáiz, J., Tudor, L., Uzun, S., Kozumplik, O., Svob Strac, D., Zarkovic, N., & Pivac, N. (2021). Metabolomics in posttraumatic stress disorder: Untargeted metabolomic analysis of plasma samples from Croatian war veterans. Free Radical Biology and Medicine, 162, 636–641. 10.1016/j.freeradbiomed.2020.11.024

Konjevod, M., Sáiz, J., Nikolac Perkovic, M., Nedic Erjavec, G., Tudor, L., Uzun, S., Kozumplik, O., Barbas, C., Zarkovic, N., Pivac, N., & Strac, D. S. (2022). Plasma lipidomics in subjects with combat posttraumatic stress disorder. Free Radical Biology and Medicine, 189, 169–177. 10.1016/j.freeradbiomed.2022.07.012

Kuan, P.-F., Yang, X., Kotov, R., Clouston, S., Bromet, E., & Luft, B. J. (2022). Metabolomics analysis of post-traumatic stress disorder symptoms in World Trade Center responders. Translational Psychiatry, 12(1), Article 1. 10.1038/s41398-022-01940-y

Lambert, L. E., French, J. F., Whitten, J. P., Baron, B. M., & McDonald, I. A. (1992). Characterization of cell selectivity of two novel inhibitors of nitric oxide synthesis. European Journal of Pharmacology, 216(1), 131–134. 10.1016/0014-2999(92)90221-O

Leitinger, N. (2003). Oxidized phospholipids as modulators of inflammation in atherosclerosis: Current Opinion in Lipidology, 14(5), 421–430. 10.1097/00041433-200310000-00002

Levine, A. B., Levine, L. M., & Levine, T. B. (2013). Posttraumatic Stress Disorder and Cardiometabolic Disease. Cardiology, 127(1), 1–19. 10.1159/000354910

Levine, G. N., Cohen, B. E., Commodore-Mensah, Y., Fleury, J., Huffman, J. C., Khalid, U., Labarthe, D. R., Lavretsky, H., Michos, E. D., Spatz, E. S., Kubzansky, L. D., & null, null. (2021). Psychological Health, Well-Being, and the Mind-Heart-Body Connection: A Scientific Statement From the American Heart Association. Circulation, 143(10), e763– e783. 10.1161/CIR.0000000000000947

Li, D., Misialek, J. R., Jack, C. R., Mielke, M. M., Knopman, D., Gottesman, R., Mosley, T., & Alonso, A. (2019). Plasma Metabolites Associated with Brain MRI Measures of Neurodegeneration in Older Adults in the Atherosclerosis Risk in Communities– Neurocognitive Study (ARIC-NCS). International Journal of Molecular Sciences, 20(7), Article 7. 10.3390/ijms20071744

Li, J., Guasch-Ferré, M., Chung, W., Ruiz-Canela, M., Toledo, E., Corella, D., Bhupathiraju, S. N., Tobias, D. K., Tabung, F. K., Hu, J., Zhao, T., Turman, C., Feng, Y.-C. A., Clish, C. B., Mucci, L., Eliassen, A. H., Costenbader, K. H., Karlson, E. W., Wolpin, B. M.,…Liang, L. (2020). The Mediterranean diet, plasma metabolome, and cardiovascular disease risk. European Heart Journal, 41(28), 2645–2656. 10.1093/eurheartj/ehaa209

Lin, P.-Y., Huang, S.-Y., & Su, K.-P. (2010). A Meta-Analytic Review of Polyunsaturated Fatty Acid Compositions in Patients with Depression. Biological Psychiatry, 68(2), 140–147. 10.1016/j.biopsych.2010.03.018

Meikle, P. J., & Summers, S. A. (2017). Sphingolipids and phospholipids in insulin resistance and related metabolic disorders. Nature Reviews Endocrinology, 13(2), Article 2. 10.1038/nrendo.2016.169

Mellon, S. H., Bersani, F. S., Lindqvist, D., Hammamieh, R., Donohue, D., Dean, K., Jett, M., Yehuda, R., Flory, J., Reus, V. I., Bierer, L. M., Makotkine, I., Abu Amara, D., Henn Haase, C., Coy, M., Doyle, F. J., Marmar, C., & Wolkowitz, O. M. (2019). Metabolomic analysis of male combat veterans with post traumatic stress disorder. PLOS ONE, 14(3), e0213839. 10.1371/journal.pone.0213839

Mellon, S. H., Gautam, A., Hammamieh, R., Jett, M., & Wolkowitz, O. M. (2018). Metabolism, Metabolomics, and Inflammation in Posttraumatic Stress Disorder. Biological Psychiatry, 83(10), 866–875. 10.1016/j.biopsych.2018.02.007

Michopoulos, V., Vester, A., & Neigh, G. (2016). Posttraumatic stress disorder: A metabolic disorder in disguise? Experimental Neurology, 284, 220–229. 10.1016/j.expneurol.2016.05.038

Miller, G. E., Chen, E., & Parker, K. J. (2011). Psychological stress in childhood and susceptibility to the chronic diseases of aging: Moving toward a model of behavioral and biological mechanisms. Psychological Bulletin, 137(6), 959–997. 10.1037/a0024768

O’Donovan, A., Ahmadian, A. J., Neylan, T. C., Pacult, M. A., Edmondson, D., & Cohen, B. E. (2017). Current posttraumatic stress disorder and exaggerated threat sensitivity associated with elevated inflammation in the Mind Your Heart Study. Brain, Behavior, and Immunity, 60, 198–205. 10.1016/j.bbi.2016.10.014

Olff, M. (2017). Sex and gender differences in post-traumatic stress disorder: An update. European Journal of Psychotraumatology, 8(sup4), 1351204. 10.1080/20008198.2017.1351204

Oosthuizen, F., Wegener, G., & Harvey, B. H. (2005). Nitric oxide as inflammatory mediator in post-traumatic stress disorder (PTSD): Evidence from an animal model. Neuropsychiatric Disease and Treatment, 1(2), 109–123. 10.2147/nedt.S61049

Pascual Cuadrado, D., Todorov, H., Lerner, R., Islami, L., Bindila, L., Gerber, S., & Lutz, B. (2022). Long-term molecular differences between resilient and susceptible mice after a single traumatic exposure. British Journal of Pharmacology, 179(17), 4161–4180. 10.1111/bph.15697

Patti, G. J., Yanes, O., & Siuzdak, G. (2012). Metabolomics: The apogee of the omics trilogy. Nature Reviews Molecular Cell Biology, 13(4), Article 4. 10.1038/nrm3314

Paynter, N. P., Balasubramanian, R., Giulianini, F., Wang, D. D., Tinker, L. F., Gopal, S., Deik, A. A., Bullock, K., Pierce, K. A., Scott, J., Martínez-González, M. A., Estruch, R., Manson, J. E., Cook, N. R., Albert, C. M., Clish, C. B., & Rexrode, K. M. (2018). Metabolic Predictors of Incident Coronary Heart Disease in Women. Circulation, 137(8), 841–853. 10.1161/CIRCULATIONAHA.117.029468

Pinto, J. V., Moulin, T. C., & Amaral, O. B. (2017). On the transdiagnostic nature of peripheral biomarkers in major psychiatric disorders: A systematic review. Neuroscience & Biobehavioral Reviews, 83, 97–108. 10.1016/j.neubiorev.2017.10.001

Ploski, J. E., & Vaidya, V. A. (2021). The Neurocircuitry of Posttraumatic Stress Disorder and Major Depression: Insights Into Overlapping and Distinct Circuit Dysfunction—A Tribute to Ron Duman. Biological Psychiatry, 90(2), 109–117. 10.1016/j.biopsych.2021.04.009

Pu, J., Liu, Y., Zhang, H., Tian, L., Gui, S., Yu, Y., Chen, X., Chen, Y., Yang, L., Ran, Y., Zhong, X., Xu, S., Song, X., Liu, L., Zheng, P., Wang, H., & Xie, P. (2020). An integrated meta-analysis of peripheral blood metabolites and biological functions in major depressive disorder. Molecular Psychiatry, 1–12. 10.1038/s41380-020-0645-4

Ratanatharathorn, A., Roberts, A. L., Chibnik, L. B., Choi, K. W., De Vivo, I., Kim, Y., Nishimi, K., Rimm, E. B., Sumner, J. A., Kubzansky, L. D., & Koenen, K. C. (2022). Posttraumatic Stress Disorder, Depression, and Accelerated Aging: Leukocyte Telomere Length in the Nurses’ Health Study II. Biological Psychiatry Global Open Science. 10.1016/j.bpsgos.2022.05.006

Roberts, A. L., Huang, T., Koenen, K. C., Kim, Y., Kubzansky, L. D., & Tworoger, S. S. (2019). Posttraumatic Stress Disorder Is Associated with Increased Risk of Ovarian Cancer: A Prospective and Retrospective Longitudinal Cohort Study. Cancer Research, 79(19), 5113–5120. 10.1158/0008-5472.CAN-19-1222

Roberts, A. L., Kubzansky, L. D., Chibnik, L. B., Rimm, E. B., & Koenen, K. C. (2020). Association of Posttraumatic Stress and Depressive Symptoms With Mortality in Women. JAMA Network Open, 3(12), e2027935. 10.1001/jamanetworkopen.2020.27935

Schnurr, P. P., Friedman, M. J., & Bernardy, N. C. (2002). Research on posttraumatic stress disorder: Epidemiology, pathophysiology, and assessment. Journal of Clinical Psychology, 58(8), 877–889. 10.1002/jclp.10064

Shutta, K. H., Balasubramanian, R., Huang, T., Jha, S. C., Zeleznik, O. A., Kroenke, C. H., Tinker, L. F., Smoller, J. W., Casanova, R., Tworoger, S. S., Manson, J. E., Clish, C. B., Rexrode, K. M., Hankinson, S. E., & Kubzansky, L. D. (2021). Plasma metabolomic profiles associated with chronic distress in women. Psychoneuroendocrinology, 133, 105420. 10.1016/j.psyneuen.2021.105420

Sumner, J. A., Kubzansky, L. D., Elkind, M. S. V., Roberts, A. L., Agnew-Blais, J., Chen, Q., Cerdá, M., Rexrode, K. M., Rich-Edwards, J. W., Spiegelman, D., Suglia, S. F., Rimm, E. B., & Koenen, K. C. (2015). Trauma Exposure and Posttraumatic Stress Disorder Symptoms Predict Onset of Cardiovascular Events in Women. Circulation, 132(4), 251–259. 10.1161/CIRCULATIONAHA.114.014492

Sumner, J. A., Nishimi, K. M., Koenen, K. C., Roberts, A. L., & Kubzansky, L. D. (2020). Posttraumatic Stress Disorder and Inflammation: Untangling Issues of Bidirectionality. Biological Psychiatry, 87(10), 885–897. 10.1016/j.biopsych.2019.11.005

van der Spek, A., Stewart, I. D., Kühnel, B., Pietzner, M., Alshehri, T., Gauß, F., Hysi, P. G., MahmoudianDehkordi, S., Heinken, A., Luik, A. I., Ladwig, K.-H., Kastenmüller, G., Menni, C., Hertel, J., Ikram, M. A., de Mutsert, R., Suhre, K., Gieger, C., Strauch, K.,…Amin, N. (2023). Circulating metabolites modulated by diet are associated with depression. Molecular Psychiatry, 1–14. 10.1038/s41380-023-02180-2

Venn-Watson, S., Reiner, J., & Jensen, E. D. (2022). Pentadecanoylcarnitine is a newly discovered endocannabinoid with pleiotropic activities relevant to supporting physical and mental health. Scientific Reports, 12(1), Article 1. 10.1038/s41598-022-18266-w

Wang, F., Tessier, A.-J., Liang, L., Wittenbecher, C., Haslam, D. E., Fernández-Duval, G., Heather Eliassen, A., Rexrode, K. M., Tobias, D. K., Li, J., Zeleznik, O., Grodstein, F., Martínez-González, M. A., Salas-Salvadó, J., Clish, C., Lee, K. H., Sun, Q., Stampfer, M. J., Hu, F. B., & Guasch-Ferré, M. (2023). Plasma metabolomic profiles associated with mortality and longevity in a prospective analysis of 13,512 individuals. Nature Communications, 14(1), Article 1. 10.1038/s41467-023-41515-z

Waszczuk, M. A., Jonas, K. G., Bornovalova, M., Breen, G., Bulik, C. M., Docherty, A. R., Eley, T. C., Hettema, J. M., Kotov, R., Krueger, R. F., Lencz, T., Li, J. J., Vassos, E., & Waldman, I. D. (2023). Dimensional and transdiagnostic phenotypes in psychiatric genome-wide association studies. Molecular Psychiatry, 1–11. 10.1038/s41380-023-02142-8

Wishart, D. S., Guo, A., Oler, E., Wang, F., Anjum, A., Peters, H., Dizon, R., Sayeeda, Z., Tian, S., Lee, B. L., Berjanskii, M., Mah, R., Yamamoto, M., Jovel, J., Torres-Calzada, C., Hiebert-Giesbrecht, M., Lui, V. W., Varshavi, D., Varshavi, D.,…Gautam, V. (2022). HMDB 5.0: The Human Metabolome Database for 2022. Nucleic Acids Research, 50(D1), D622–D631. 10.1093/nar/gkab1062

Yehuda, R. (2001). Biology of posttraumatic stress disorder. The Journal of Clinical Psychiatry, 62 Suppl 17, 41–46.

Zeleznik, O. A., Huang, T., Patel, C. J., Poole, E. M., Clish, C. B., Armaiz-Pena, G. N., Nagaraja, A. S., Eliassen, A. H., Shutta, K. H., Balasubramanian, R., Kubzansky, L. D., Hankinson, S. E., Sood, A. K., & Tworoger, S. S. (2022). Plasma and ovarian metabolomic responses to chronic stress in female mice (p. 2022.01.03.474852). bioRxiv. 10.1101/2022.01.03.474852

Zhu, Y., Jha, S. C., Shutta, K. H., Huang, T., Balasubramanian, R., Clish, C. B., Hankinson, S. E., & Kubzansky, L. D. (2022). Psychological distress and metabolomic markers: A systematic review of posttraumatic stress disorder, anxiety, and subclinical distress. Neuroscience & Biobehavioral Reviews, 143, 104954. 10.1016/j.neubiorev.2022.104954

