## Supplement for "Persistent PTSD symptoms are associated with plasma metabolic alterations relevant to long-term health: A metabolome-wide investigation in women"

**Supplementary Methods**

*NHSII blood collection procedure*

NHSII participants who responded to the 1995 biennial survey and had not reported a cancer diagnosis were considered eligible for participation in the first blood collection in 1996. A total of 92,888 eligible women were invited. If the participant was premenopausal at the time of blood collection, she was instructed to collect two samples, timed within the follicular and mid-luteal phases of the menstrual cycle (Missmer et al., 2006). Blood samples collected during the mid-luteal phase were used in metabolomic profiling. Other participants provided an untimed, single blood sample. All participants completed a questionnaire about variables related to the blood collection process at the time of the collection.

Out of the 92,888 eligible participants, a total of 29,611 (32%) women provided blood samples (N=18,521 in the timed collection). Women who participated in the first blood collection were invited to provide a second sample between 2010 and 2012, and 16,510 (56%) women returned a second sample which followed essentially identical collection, processing, and storage procedures.

At all blood collection time points, participants arranged to have their blood samples collected (in heparin tubes that we provided) and mailed to the NHSII laboratory overnight to be processed into plasma, red blood cells, and white blood cells. Blood samples were separated into small aliquots and stored in the vapor phase of liquid nitrogen freezers (-130°C or lower).

*Overview of analytic subsamples*

Our analytic sample comes from three independent subsamples with non-overlapping participants, all nested within the NHSII cohort (**Supplemental Figure S1**).

**First,** eight sub-studies ranging in size from 112 to 2089 women have obtained metabolomic data (primarily leveraging samples from a blood collection between 1996-1999) to assess associations between plasma metabolomics and several health conditions, including breast cancer, diabetes, inflammatory bowel disease, ovarian cancer, primary open angle glaucoma, rheumatoid arthritis, stroke, and stress. Drawing on recent cohort-wide efforts to harmonize metabolomics data across sub-studies (hereby referred to as the *NHSII merged dataset*), we identified women who completed the PTSD screener administered in 2008 and had harmonized metabolomic data. With the exception of the stress sub-study, the sub-studies followed a prospective nested case-control design; participants diagnosed with conditions assessed in the sub-studies within two years before or after blood collections were excluded from our analytic sample, as these conditions could contribute to both PTSD symptoms and metabolic alterations. Among the women in the NHSII merged dataset, 2276 women only provided a blood sample between 1996-1999, 44 women only provided a blood sample between 2010-2012, and 115 women provided blood samples at both time points. As such, the final analytic sample from the NHSII merged dataset includes data from 2435 women and 2550 samples in total. Of note, for the 115 women with two blood samples included in the analyses, the dependence between samples within individual was accounted for in the subsequent analyses using generalized estimating equations with exchangeable covariance structure.

**Second,** PTSD and metabolomics data were assessed in the Mind-Body Study (MBS; n=204), a sub-study designed to investigate connections between psychosocial factors and biological processes (Huang et al., 2019). The blood collection took place between 2013 and 2014.

**Third,** we generated new metabolomic data for the current investigation in a subsample hereafter considered the *Severe Distress Sample*. For this subsample we selected 196 women who contributed blood samples between 2010 and 2012 to enrich our study with more women who reported severe PTSD symptoms. The selection criteria are described in the next section.

Overall, >80% of the sample came from the NHSII merged dataset with metabolomic assays obtained at the blood collection between 1996-1999, for which participants were on average 44.6 years of age and predominantly pre-menopausal (73.9%). Characteristics of women in the NHSII merged dataset, MBS, and Severe Distress Sample whose metabolomic assays came from the blood collections between 2010-2012 or 2013-2014, were similar: the average age was 56.8 to 60.5 years, and >70% of the women were post-menopausal. Health-related factors were differentially distributed between the subsamples following an age-associated pattern, with higher prevalence of hypertension and type 2 diabetes history in the subsamples with older participants.

*Selection criteria for the Severe Distress Sample*

The Severe Distress Sample was designed to facilitate a high contrast comparison of women experiencing more severe PTSD with those who had not experienced traumatic events and exhibited low levels of psychological distress. Women for this subsample were drawn from those who completed the NHSII PTSD questionnaire administered in 2008. Briefly, in 2008, active participants who responded to the 2007 biennial survey were invited to complete a self-report screening questionnaire assessing lifetime exposure to traumatic events and PTSD. Out of the 50,953 respondents, a subset of 2,112 probable PTSD cases and 2,001 probable controls were randomly selected to participate in a phone-based semi-structured diagnostic interview. To be eligible for inclusion in the selected sample, participants must: 1) have completed the PTSD sub-study in 2008 (for women identified as having high symptoms, completion of the phone-based clinical interview is also required to validate their PTSD status); 2) have provided a blood sample from the 2010-2012 NHSII blood collection, the collection period closest in time to PTSD assessment (within 2-4 years); 3) be free of cancer or cardiovascular conditions (myocardial infarction, stroke, or having undergone a coronary artery bypass grafting procedure) at the time of blood collection; and 4) self-identify as non-Hispanic Caucasian in order to minimize other sources of heterogeneity due to race and ethnic differences. Investigators selected 100 women who had a confirmed diagnosis of PTSD based on DSM-IV criteria and were considered cases. They also selected 100 women to serve in the comparison group with low exposure to distress. These women were selected on the basis of 1) either never reporting exposure to traumatic events or reporting exposure to only one traumatic event (excluding child death, physical assault, sexual abuse, or other) and not endorsing any PTSD symptoms, and 2) did not have depression (based on not meeting cut points for probable depression using a measure of depressive symptoms assessed in 2008 with the 10-item Center of Epidemiologic Studies Depression Scale (Björgvinsson et al., 2013), antidepressant use, and self-reported physician diagnosis of major depressive disorder assessed in 2009) or anxiety (assessed based on not meeting a previously established cut-point for probable anxiety on the Crown Crisp Index (Crown & Crisp, 1966) administered in in 2005).

Of note, this sample of 200 women was originally constructed as a case-control sample (severe distress vs not severely distressed) and the pairs were matched on age, menopausal status, fasting status, and time of blood draw. However, to harmonize the analytic setup with other subsamples used in the current study and maximize the total sample size, we did not analyze this dataset as a matched sample; rather, we adjusted for all matching factors in the baseline model. In addition, to measure PTSD consistently with the other subsamples, PTSD data collected in the 2008 questionnaire were used to construct measures of exposures, following the same analytic pipeline as the NHSII merged dataset and MBS subsamples. At the analysis phase, four samples from four women were additionally removed because they were already in the NHSII merged dataset. The final sample size for the Severe Distress Sample was 196.

*Metabolite profiling and quality control*

Plasma metabolomic profiling was performed at the Broad Institute of the Massachusetts Institute of Technology and Harvard using a platform comprising complementary liquid chromatography tandem mass spectrometry (LC-MS)-based methods: HILIC-pos and HILIC-neg for water soluble metabolites (e.g., amino acids), and C8-pos for polar and nonpolar lipid metabolites. Duplicated metabolites measured in more than one LC-MS method were removed based on guidance provided by the Broad Institute. Additionally, 89 metabolites did not pass the processing method pilot (i.e., the usual delay between the blood collection and processing substantially affected the assay results) and were removed from subsequent analyses.

339 metabolites were available in all subsamples and retained in our analyses. Blinded replicates were added to every laboratory batch to assess reproducibility; the median of the percent coefficient of variation (CV) of the analyzed metabolites (10th - 90th percentile) was 11.6 (7.8-24.6) from the 1996-1999 blood collection in the NHSII merged dataset, 12.1 (8.2-25.2) from the 2010-2012 blood collection of the NHSII merged dataset, 9.0 (5.3-24.2) in MBS, and 9.8 (5.9-16.8) in the Severe Distress Sample.

As previously reported, plasma metabolites measured in the NHSII had reasonable long-term within-woman stability (Zeleznik et al., 2022). Among 336 metabolites with available 10-year within-person intraclass correlation (ICC), the median ICC was 0.6 (10th - 90th percentile range: 0.3-0.8).

*MDS development and construction*

We examined a previously defined MDS capturing metabolic alterations linked to depression and anxiety in relation to our PTSD measures. The process of developing and optimizing the MDS has been described in detail by Balasubramanian et al. ((2023). Briefly, the MDS was developed in a 1:1 matched case-control sample of chronic distress (defined based on repeated assessments of depression and anxiety prior to blood sampling; n=558) nested within the Nurses’ Health Study I, another prospective cohort study of women similar to the Nurses’ Health Study II with non-overlapping samples. In other words, there are no women in this case-control sample of chronic distress who are also in the three subsamples in the present study’s analyses. The authors selected 100 unique metabolites based on prior evidence of metabolomic associations with distress in the same underlying population, and performed variable selection using LASSO conditional logistic regression, which resulted in a set of 20 metabolites. Metabolite weights were estimated in this case-control sample of distress using unpenalized conditional logistic regression. The MDS is a linear combination of log transformed and standardized metabolites as shown below:

$MDS=\sum_{j=1}^{20} \beta_{j}x_{j}$.

The regression coefficients ($\beta_{j}$) in the MDS were estimated in the matched case-control dataset of chronic distress mentioned above (N=558) in a conditional logistic regression model.

Subsequent analysis by Balasubramanian et al. revealed that higher scores on this MDS were associated with increased incident coronary heart disease risk and cardiovascular disease risk in independent validation samples of both men and women, as well as incident diabetes in a case-control sample of women (Huang et al., 2023).

Out of the 20 metabolites, 19 were available in our study. See **Supplementary Table S4** for a list of the metabolites in the MDS and their specific weights. The omission of one metabolite (arachidonate) was due to differences in the designs of the original study deriving the MDS and our current study. In our prior work developing the MDS, metabolomic profiling was performed on all four platforms available at the Broad Institute: HILIC-pos and HILIC-neg for water soluble metabolites (e.g., amino acids), C8-pos for polar and nonpolar lipid metabolites, and C18-neg for free fatty acids and bile acids. In the current study, we restricted the analysis to metabolites measured in all three datasets and not all platforms were available in all; metabolites measured only by the C18-neg method were not available, including arachidonate. In the original MDS study samples, the revised version of the MDS comprised of 19 metabolites (i.e., the measure in our study) was highly correlated with the full 20-metabolite score (*r*=0.97) (Huang et al., 2023).

Because of varying sample sizes for different metabolites and platforms available across the three subsamples, we restricted the MDS analysis to N=2006 participants with complete data on all 19 metabolites in the current study.

*Covariate measures*

Covariates included relevant demographic factors, medical conditions, and biobehavioral factors that could confound or lie on the pathway between PTSD and metabolite concentrations. Age, race/ethnicity, and height were reported in the baseline survey at study recruitment in 1989. Participants’ menopausal status (pre-menopausal, post-menopausal, or missing/unknown) and fasting status at blood draw (fasting >= 8 hours, non-fasting or unknown) were self-reported on the questionnaire returned with the blood collection kit at each blood collection. Use of statin or other lipid lowering medication was queried (yes/no) on each biennial survey starting in 1999; for the first, second, and MBS blood collections, measures from 1999, 2009, and the MBS sub-study questionnaires, respectively, were used. Hormone therapy use was measured in the questionnaire administered at the time of each blood collections. Clinical diagnoses of hypertension and type 2 diabetes were self-reported (yes/no) at each biennial survey, and the participant was considered to have a history of hypertension or type 2 diabetes if the reported year of diagnosis preceded the blood collection date.

Biobehavioral factors included measures of diet, caffeine intake, and alcohol consumption, which were derived from the validated semi-quantitative Food Frequency Questionnaire (FFQ) (Willett et al., 1985) administered in 1999, 2007, and 2011 biennial surveys. These surveys are the waves closest in time to the three blood collections considered in our analyses, respectively. We derived a measure overall diet quality, the validated Alternate Healthy Eating Index (AHEI), following prior work whereby consumption levels of each food item are combined to provide a continuous score, with higher scores indicating a healthier diet (Chiuve et al., 2012). Caffeine intake (mg/day) and alcohol consumption (g/day) were also coded based on assessments of individual food intake in accordance with established cohort conventions, which has been validated in prior studies (Salvini et al., 1989; Glovannucci et al., 1991). Also following prior work in the cohort, physical activity was measured by metabolic equivalent of energy (MET) hours per week (Ainsworth et al., 1993), based on participants’ reports of their leisure-time activities in the biennial surveys. Activity levels were derived from the survey taken closest in time to the blood collection from which participants’ metabolite profiles were obtained (1997 for the 1996-1999 blood collection; 2009 were used for the 2010-2012 and 2013-2014 blood collections). Smoking status and frequency were reported at the time of each blood draw. Body mass index (BMI; kg/ m^2^) was derived from self-reported height taken at baseline and weight reported at blood draw; this self-report based measure has been validated against objective measures in the Nurses’ Health Study (Rimm et al., 1990). Of note, for the 115 participants with repeated blood samples available from two time points, covariate data were updated to match the time of blood collection.

Of note, while biobehavioral covariates can also serve as mechanisms underlying the association between PTSD and metabolite markers, we included covariate measures from a single time point and could not assess potential mediation.

*Regression model interpretation*

In both the analysis of individual metabolites and the MDS, we included all three exposure variables in each model simultaneously:

$$E\left[ \mathrm{metabolite}_{j} \right]= \beta_{0}+\beta_{1}Persistent PTSD Sx Count+ \beta_{2} Remitted PTSD+ \beta_{3}Trauma+ \boldsymbol{\beta}_{\mathbf{4}} \mathbf{Covariates}$$

The model allows us to disentangle associations pertaining to different sources of exposure. Specifically, $\beta_{1}$captures the difference in the *j^th^* metabolite associated with having one additional persistent PTSD symptom, among trauma-exposed participants whose PTSD did not remit at the time of survey administration. $\beta_{2}$captures the difference in the *j^th^* metabolite level comparing trauma-exposed participants with remitted lifetime PTSD to trauma-exposed participants who did not meet lifetime diagnostic criteria for PTSD. $\beta_{3}$ captures the difference in the *j^h^* metabolite level comparing trauma exposed participants who never had PTSD to participants unexposed to trauma at the time of blood collection. All estimates were conditional on additional covariates described above and in the main text.

*Additional detail on the main statistical approach*

**First,** we assessed whether levels of metabolites previously linked to depression and anxiety were associated with PTSD and trauma exposures. Specifically, we pursued two sets of analyses: (a) testing associations with the MDS; and (b) testing associations with each of the 19 available metabolites in the MDS. To estimate associations between PTSD/trauma exposures and the MDS, we first performed separate analyses within each dataset due to the heterogeneity in sample selection procedures, timing of metabolite profiling, and covariate measurements. In the NHSII merged dataset, we fitted a generalized estimating equation with exchangeable covariance structure to account for within-person dependence of the 115 repeated samples available at two time points. In the MBS and Severe Distress Sample with only one blood collection each, linear regression models were used. We accounted for the differences in age, fasting status, and menopausal status at the three blood collections by including these factors as covariates in all models. We then combined the three subsample-specific estimates using a random effects meta-analysis, which allows the effects (i.e., associations between PTSD/trauma exposures and the MDS) to vary in each subsample, rather than assuming the same true effect across subsamples. We further calculated Cochran’s Q-statistics to assess between-study heterogeneity. To test associations of each metabolite in the MDS with PTSD/trauma exposures, we conducted random effects meta-analysis for each of 19 models with individual metabolite levels as the dependent variables and followed the same modeling strategy used for the MDS.

In our **second, broader analysis**, we performed a metabolome-wide agnostic analysis to identify metabolic markers associated with PTSD and trauma exposures. Extending the metabolite-level analysis described above to all 339 available metabolites (including the 19 metabolites that comprised the MDS), we obtained meta-analyzed estimates for the association between each metabolite and PTSD/trauma exposure. Of note, the overall sample size varied by metabolite in the agnostic analyses: not all metabolites were measured or detected in all sub-studies within the NHSII merged dataset due to different laboratory and quality control processes. As such, the analytic sample sizes for 339 metabolites across the three substudies ranged from 438 to 2779 unique individuals, with 83% of the metabolites available for analysis in at least 2000 individuals.

*Differential network analysis of persistent symptoms*

To follow up on the results obtained from the agnostic analysis and characterize systems-level metabolic differences linked to PTSD, we constructed a differential network comparing participants who reported any persistent PTSD symptoms (exposed) and participants who had no persistent symptoms (unexposed; including both participants with remitted symptoms or reporting no trauma exposure) using the DINGO algorithm (Ha et al., 2015) with data from all three subsamples. For this analysis, we included the 29 metabolites that were associated with persistent PTSD symptoms in the meta-analysis of the minimally adjusted model. We chose to use metabolites from this model to strike the balance between including a wide range of metabolites with potentially meaningful signals while also retaining adequate sample size (including all 339 metabolites would lead to high levels of noise and reduce our sample size; performing the analyses with only the nine metabolites identified in Model 2 could result in missing important signals that may not have come up due to power). Thus, we used the set of metabolites that were retained in the initial models. Following this strategy, complete metabolite data were available from 858 independent participants across the three subsamples. For women with two blood samples available in the agnostic analyses, only data from the blood collection between 2010-2012 were retained to prioritize information collected closer to the PTSD assessment.

After performing the DINGO analysis, we created thresholds for the resulting differential network to include only edges that significantly differed between exposed and unexposed participants (FDR < 0.05). Within this differential network, we calculated three types of network centrality measures: hub score, betweenness, and closeness. These measures were presented to aid our interpretations of any given metabolite’s relative importance in driving differences between being exposed versus unexposed to persistent PTSD symptoms. Specifically, hub centrality captures the relative influence of a node in the network (Kleinberg, 1999), betweenness centrality is based on the number of shortest paths through the network that include the node (Brandes, 2001; Freeman, 1978); and closeness centrality is based on the average distance from each node to all the other nodes in the network (Freeman, 1978). Betweenness and closeness centrality treat edge weights as a measure of distance between nodes; because our differential edges correspond to differences in partial correlations, when calculating the network measures, we converted the weights by taking the reciprocal of their absolute values, so that low edge weights correspond to high differences in partial correlations and high edge weights correspond to low differences in partial correlations. Thus, betweenness and closeness centrality measures are based on path lengths that are shorter for paths linking differential edges and longer for paths that do not appear different between exposed and unexposed individuals.

*Metabolite class enrichment analysis*

Assessing whether the overall patterns of associations with PTSD versus with depression or anxiety may differ at the metabolite class level rather than restricting the analyses to metabolites in the MDS, we performed metabolite set enrichment analysis using the gene set enrichment analysis algorithm implemented in the *fgsea* package (Korotkevich et al., 2021). Briefly, to evaluate the enrichment of metabolite classes for associations with PTSD, 335 metabolites that were included in the PTSD agnostic analyses and in previously reported agnostic analyses with depression/anxiety were first annotated to a unique metabolite class using data from the Human Metabolome Database (Wishart et al., 2022). **First,** test statistics obtained from the minimally adjusted meta-analysis that correspond to the association between persistent PTSD symptoms and each metabolite were ranked from the highest positive value to the lowest negative value. Normalized enrichment scores were calculated to represent the degree to which metabolites from each class may be enriched at the top (i.e., more likely to be positively associated) or bottom (i.e., more likely to be negatively associated) of the ranked list. An adaptive multilevel splitting Monte Carlo approach was used, as implemented by the *fgseaMultilevel* function, with 1000 permutations. Metabolite classes with at least 5 metabolites annotated to that class were considered based on prior recommendations (Korotkevich et al., 2021; Sergushichev, 2016). We then applied false discovery correction (FDR p<0.05) to these enrichment scores.

**Second,** to evaluate the enrichment of metabolite classes for associations with depression or anxiety, we repeated the metabolite set enrichment analysis described above with test statistics reported by Shutta et al. as input (Shutta et al., 2021, p. 202). The same procedure was followed to calculate the normalized enrichment scores. Analysis was restricted to metabolites that were also available in our study. We compared results from the two sets of enrichment analyses as illustrated in **Supplemental Figure S2.**

**Sensitivity Analysis**

The primary results from the agnostic analysis were obtained by analyzing available data from participants who did not receive a diagnosis of the conditions or diseases (e.g., breast cancer) assessed in the sub-studies included within the NHSII merged dataset within two years before or after blood collection; those who did receive a diagnosis in that time period were excluded, as disease onset and diagnoses could cause substantial changes in biobehavioral states that may contribute to both PTSD symptoms and metabolomic alterations. As a sensitivity analysis, we also tested whether the observed associations still held when we restricted the NHSII merged dataset to individuals who *never* developed the disease in the original case-control substudies with various physical health endpoints (i.e., the controls) as well as samples from the stress sub-study (which was the only sub-study not following a case-control design). Data were available from 1944 samples.

Restricting samples from the NHSII merged dataset to controls and meta-analyzing with the MBS and Severe Distress Sample following the same procedure as in the primary analyses, we found that all nine metabolites significant in Model 2 of the primary analysis were nominally significant and five remained significant after metabolome-wide FDR corrections for 339 metabolites (**Supplemental Figure S3**). Point estimates and confidence intervals were similar to those identified in the primary analysis.

**References**

Ainsworth, B. E., Haskell, W. L., Leon, A. S., Jacobs, D. R., Montoye, H. J., Sallis, J. F., & Paffenbarger, R. S. (1993). Compendium of physical activities: Classification of energy costs of human physical activities. *Medicine and Science in Sports and Exercise*, *25*(1), 71–80. https://doi.org/10.1249/00005768-199301000-00011

Balasubramanian, R., Shutta, K. H., Guasch-Ferre, M., Huang, T., Jha, S. C., Zhu, Y., Shadyab, A. H., Manson, J. E., Corella, D., Fitó, M., Hu, F. B., Rexrode, K. M., Clish, C. B., Hankinson, S. E., & Kubzansky, L. D. (2023). Metabolomic profiles of chronic distress are associated with cardiovascular disease risk and inflammation-related risk factors. *Brain, Behavior, and Immunity*. https://doi.org/10.1016/j.bbi.2023.08.010

Björgvinsson, T., Kertz, S. J., Bigda-Peyton, J. S., McCoy, K. L., & Aderka, I. M. (2013). Psychometric Properties of the CES-D-10 in a Psychiatric Sample. *Assessment*, *20*(4), 429–436. https://doi.org/10.1177/1073191113481998

Brandes, U. (2001). A faster algorithm for betweenness centrality*. *The Journal of Mathematical Sociology*, *25*(2), 163–177. https://doi.org/10.1080/0022250X.2001.9990249

Chiuve, S. E., Fung, T. T., Rimm, E. B., Hu, F. B., McCullough, M. L., Wang, M., Stampfer, M. J., & Willett, W. C. (2012). Alternative Dietary Indices Both Strongly Predict Risk of Chronic Disease, ,. *The Journal of Nutrition*, *142*(6), 1009–1018. https://doi.org/10.3945/jn.111.157222

Crown, S., & Crisp, A. H. (1966). A short clinical diagnostic self-rating scale for psychoneurotic patients. The Middlesex Hospital Questionnaire (M.H.Q.). *The British Journal of Psychiatry: The Journal of Mental Science*, *112*(490), 917–923. https://doi.org/10.1192/bjp.112.490.917

Freeman, L. C. (1978). Centrality in social networks conceptual clarification. *Social Networks*, *1*(3), 215–239. https://doi.org/10.1016/0378-8733(78)90021-7

Glovannucci, E., Colditz, G., Stampfer, M. J., Rimm, E. B., Litin, L., Sampson, L., & Willett, W. C. (1991). The Assessment of Alcohol Consumption by a Simple Self-administered Questionnaire. *American Journal of Epidemiology*, *133*(8), 810–817. https://doi.org/10.1093/oxfordjournals.aje.a115960

Ha, M. J., Baladandayuthapani, V., & Do, K.-A. (2015). DINGO: Differential network analysis in genomics. *Bioinformatics*, *31*(21), 3413–3420. https://doi.org/10.1093/bioinformatics/btv406

Huang, T., Trudel-Fitzgerald, C., Poole, E. M., Sawyer, S., Kubzansky, L. D., Hankinson, S. E., Okereke, O. I., & Tworoger, S. S. (2019). The Mind–Body Study: Study design and reproducibility and interrelationships of psychosocial factors in the Nurses’ Health Study II. *Cancer Causes & Control*, *30*(7), 779–790. https://doi.org/10.1007/s10552-019-01176-0

Huang, T., Zhu, Y., Shutta, K. H., Balasubramanian, R., Zeleznik, O. A., Rexrode, K. M., Clish, C. B., Sun, Q., Hu, F. B., Kubzansky, L. D., & Hankinson, S. E. (2023). A Plasma Metabolite Score Related to Psychological Distress and Diabetes Risk: A Nested Case-control Study in US Women. *The Journal of Clinical Endocrinology & Metabolism*, dgad731. https://doi.org/10.1210/clinem/dgad731

Kleinberg, J. M. (1999). Authoritative sources in a hyperlinked environment. *Journal of the ACM*, *46*(5), 604–632. https://doi.org/10.1145/324133.324140

Korotkevich, G., Sukhov, V., Budin, N., Shpak, B., Artyomov, M. N., & Sergushichev, A. (2021). *Fast gene set enrichment analysis* (p. 060012). bioRxiv. https://doi.org/10.1101/060012

Missmer, S. A., Spiegelman, D., Bertone-Johnson, E. R., Barbieri, R. L., Pollak, M. N., & Hankinson, S. E. (2006). Reproducibility of Plasma Steroid Hormones, Prolactin, and Insulin-like Growth Factor Levels among Premenopausal Women over a 2- to 3-Year Period. *Cancer Epidemiology, Biomarkers & Prevention*, *15*(5), 972–978. https://doi.org/10.1158/1055-9965.EPI-05-0848

Rimm, E. B., Stampfer, M. J., Colditz, G. A., Chute, C. G., Litin, L. B., & Willett, W. C. (1990). Validity of Self-Reported Waist and Hip Circumferences in Men and Women. *Epidemiology*, *1*(6), 466–473.

Salvini, S., Hunter, D. J., Sampson, L., Stampfer, M. J., Colditz, G. A., Rosner, B., & Willett, W. C. (1989). Food-Based Validation of a Dietary Questionnaire: The Effects of Week-to-Week Variation in Food Consumption. *International Journal of Epidemiology*, *18*(4), 858–867. https://doi.org/10.1093/ije/18.4.858

Sergushichev, A. A. (2016). *An algorithm for fast preranked gene set enrichment analysis using cumulative statistic calculation* (p. 060012). bioRxiv. https://doi.org/10.1101/060012

Shutta, K. H., Balasubramanian, R., Huang, T., Jha, S. C., Zeleznik, O. A., Kroenke, C. H., Tinker, L. F., Smoller, J. W., Casanova, R., Tworoger, S. S., Manson, J. E., Clish, C. B., Rexrode, K. M., Hankinson, S. E., & Kubzansky, L. D. (2021). Plasma metabolomic profiles associated with chronic distress in women. *Psychoneuroendocrinology*, *133*, 105420. https://doi.org/10.1016/j.psyneuen.2021.105420

WILLETT, W. C., SAMPSON, L., STAMPFER, M. J., ROSNER, B., BAIN, C., WITSCHI, J., HENNEKENS, C. H., & SPEIZER, F. E. (1985). REPRODUCIBILITY AND VALIDITY OF A SEMIQUANTITATIVE FOOD FREQUENCY QUESTIONNAIRE. *American Journal of Epidemiology*, *122*(1), 51–65. https://doi.org/10.1093/oxfordjournals.aje.a114086

Wishart, D. S., Guo, A., Oler, E., Wang, F., Anjum, A., Peters, H., Dizon, R., Sayeeda, Z., Tian, S., Lee, B. L., Berjanskii, M., Mah, R., Yamamoto, M., Jovel, J., Torres-Calzada, C., Hiebert-Giesbrecht, M., Lui, V. W., Varshavi, D., Varshavi, D., … Gautam, V. (2022). HMDB 5.0: The Human Metabolome Database for 2022. *Nucleic Acids Research*, *50*(D1), D622–D631. https://doi.org/10.1093/nar/gkab1062

Zeleznik, O. A., Wittenbecher, C., Deik, A., Jeanfavre, S., Avila-Pacheco, J., Rosner, B., Rexrode, K. M., Clish, C. B., Hu, F. B., & Eliassen, A. H. (2022). Intrapersonal Stability of Plasma Metabolomic Profiles over 10 Years among Women. *Metabolites*, *12*(5), Article 5. https://doi.org/10.3390/metabo12050372

Supplemental Table S1A. Associations between persistent PTSD symptoms and 19 metabolites previously associated with depression and anxiety included in the metabolite-based distress score, adjusting for base covariates and medical covariates, in the random-effects meta-analysis and within each subsample. Bold values indicate significant associations after correcting for testing 19 metabolites (FDR p<0.05).

| **HMDB ID** | **Metabolite** | **Metabolite Class** | **Minimal**  **β (95% CI)** | **Minimal p-value** | **Model 2**  **β (95% CI)** | **Model 2 p-value** |
| --- | --- | --- | --- | --- | --- | --- |
| *Random-Effects Meta-Analysis* | | | | | | |
| HMDB0000112 | GABA | Carboxylic acids and derivatives | -0.04 (-0.09, 0.01) | 0.1314 | -0.04 (-0.09, 0.01) | 0.1072 |
| HMDB0000167 | threonine | Carboxylic acids and derivatives | 0.01 (-0.01, 0.04) | 0.3091 | 0.01 (-0.02, 0.04) | 0.3859 |
| HMDB0000235 | thiamine | Diazines | 0.04 (0.01, 0.07) | 0.0190 | 0.03 (0, 0.07) | 0.0271 |
| HMDB0000259 | serotonin | Indoles and derivatives | **-0.05 (-0.08, -0.02)** | **0.0020** | **-0.04 (-0.07, -0.01)** | **0.0053** |
| HMDB0000562 | creatinine | Carboxylic acids and derivatives | 0 (-0.03, 0.03) | 0.8172 | 0 (-0.03, 0.03) | 0.8519 |
| HMDB0000641 | glutamine | Carboxylic acids and derivatives | 0.01 (-0.02, 0.03) | 0.7177 | 0.01 (-0.02, 0.04) | 0.6192 |
| HMDB0000714 | hippurate | Benzene and substituted derivatives | -0.01 (-0.06, 0.04) | 0.6759 | -0.01 (-0.07, 0.04) | 0.6698 |
| HMDB0000767 | pseudouridine | Nucleoside and nucleotide analogues | 0.03 (-0.02, 0.08) | 0.2501 | 0.02 (-0.01, 0.04) | 0.3060 |
| HMDB0000929 | tryptophan | Indoles and derivatives | -0.03 (-0.05, 0) | 0.0683 | -0.03 (-0.05, 0) | 0.0745 |
| HMDB0001008 | biliverdin | Tetrapyrroles and derivatives | -0.04 (-0.09, 0) | 0.0553 | -0.03 (-0.06, 0) | 0.0268 |
| HMDB0001046 | cotinine | Pyridines and derivatives | 0.03 (0, 0.06) | 0.0355 | 0.03 (0, 0.06) | 0.0353 |
| HMDB0001886 | 3-methylxanthine | Imidazopyrimidines | 0.02 (-0.01, 0.05) | 0.1357 | 0.02 (-0.01, 0.05) | 0.1420 |
| HMDB0004824 | N2,N2-dimethylguanosine | Purine nucleosides | 0.01 (-0.02, 0.03) | 0.6993 | 0 (-0.03, 0.03) | 0.9371 |
| HMDB0004949 | C16:0 Ceramide (d18:1) | Sphingolipids | 0.03 (0, 0.06) | 0.0300 | 0.05 (-0.02, 0.12) | 0.1873 |
| HMDB0005923 | N4-acetylcytidine | Pyrimidine nucleosides | 0.03 (0, 0.05) | 0.0846 | 0.02 (-0.01, 0.05) | 0.2105 |
| HMDB0008006* | C34:3 PC | Glycerophospholipids | **0.08 (0.03, 0.13)** | **0.0029** | **0.08 (0.02, 0.13)** | **0.0072** |
| HMDB0008047* | C38:3 PC | Glycerophospholipids | **0.07 (0.02, 0.12)** | **0.0084** | 0.07 (0.01, 0.12) | 0.0268 |
| HMDB0011130 | C18:0 LPE | Glycerophospholipids | **0.04 (0.01, 0.07)** | **0.0037** | **0.04 (0.02, 0.07)** | **0.0030** |
| HMDB0011220* | C36:5 PC plasmalogen-B | Glycerophospholipids | -0.01 (-0.04, 0.01) | 0.3393 | -0.01 (-0.04, 0.01) | 0.3162 |
| *NHSII Merged Dataset* | | | | | | |
| HMDB0000112 | GABA | Carboxylic acids and derivatives | -0.01 (-0.04, 0.02) | 0.5335 | -0.01 (-0.05, 0.02) | 0.4270 |
| HMDB0000167 | threonine | Carboxylic acids and derivatives | 0.02 (-0.01, 0.05) | 0.2480 | 0.02 (-0.02, 0.05) | 0.3124 |
| HMDB0000235 | thiamine | Diazines | 0.03 (0, 0.07) | 0.0894 | 0.03 (-0.01, 0.07) | 0.0973 |
| HMDB0000259 | serotonin | Indoles and derivatives | -0.05 (-0.08, -0.01) | 0.0112 | -0.04 (-0.08, -0.01) | 0.0165 |
| HMDB0000562 | creatinine | Carboxylic acids and derivatives | 0 (-0.03, 0.04) | 0.9429 | 0 (-0.03, 0.04) | 0.8721 |
| HMDB0000641 | glutamine | Carboxylic acids and derivatives | 0.01 (-0.03, 0.04) | 0.7197 | 0.01 (-0.02, 0.04) | 0.5915 |
| HMDB0000714 | hippurate | Benzene and substituted derivatives | 0.01 (-0.03, 0.04) | 0.7667 | 0.01 (-0.03, 0.05) | 0.6893 |
| HMDB0000767 | pseudouridine | Nucleoside and nucleotide analogues | 0.01 (-0.02, 0.05) | 0.4551 | 0.01 (-0.02, 0.04) | 0.5257 |
| HMDB0000929 | tryptophan | Indoles and derivatives | -0.03 (-0.06, 0) | 0.0631 | -0.03 (-0.06, 0) | 0.0704 |
| HMDB0001008 | biliverdin | Tetrapyrroles and derivatives | -0.02 (-0.05, 0.01) | 0.1313 | -0.02 (-0.05, 0.01) | 0.1223 |
| HMDB0001046 | cotinine | Pyridines and derivatives | 0.02 (-0.01, 0.05) | 0.1534 | 0.02 (-0.01, 0.06) | 0.1414 |
| HMDB0001886 | 3-methylxanthine | Imidazopyrimidines | 0.03 (-0.01, 0.06) | 0.1188 | 0.03 (-0.01, 0.06) | 0.1319 |
| HMDB0004824 | N2,N2-dimethylguanosine | Purine nucleosides | 0 (-0.03, 0.04) | 0.9005 | 0 (-0.03, 0.04) | 0.9148 |
| HMDB0004949 | C16:0 Ceramide (d18:1) | Sphingolipids | 0.03 (0, 0.06) | 0.0900 | 0.02 (-0.01, 0.06) | 0.1261 |
| HMDB0005923 | N4-acetylcytidine | Pyrimidine nucleosides | 0.02 (-0.02, 0.05) | 0.3048 | 0.01 (-0.02, 0.04) | 0.5215 |
| HMDB0008006* | C34:3 PC | Glycerophospholipids | **0.05 (0.02, 0.08)** | **0.0019** | 0.04 (0.01, 0.07) | 0.0080 |
| HMDB0008047* | C38:3 PC | Glycerophospholipids | **0.05 (0.01, 0.08)** | **0.0046** | 0.04 (0.01, 0.07) | 0.0166 |
| HMDB0011130 | C18:0 LPE | Glycerophospholipids | **0.05 (0.01, 0.08)** | **0.0075** | 0.05 (0.01, 0.08) | 0.0073 |
| HMDB0011220* | C36:5 PC plasmalogen-B | Glycerophospholipids | -0.01 (-0.05, 0.02) | 0.4503 | -0.01 (-0.05, 0.02) | 0.3803 |
| *Mind-Body Study* | | | | | | |
| HMDB0000112 | GABA | Carboxylic acids and derivatives | -0.07 (-0.18, 0.03) | 0.1864 | -0.08 (-0.19, 0.03) | 0.1421 |
| HMDB0000167 | threonine | Carboxylic acids and derivatives | 0 (-0.1, 0.1) | 0.9859 | -0.01 (-0.12, 0.09) | 0.8238 |
| HMDB0000235 | thiamine | Diazines | 0.07 (-0.04, 0.17) | 0.2174 | 0.05 (-0.06, 0.15) | 0.3767 |
| HMDB0000259 | serotonin | Indoles and derivatives | -0.09 (-0.19, 0.02) | 0.1191 | -0.06 (-0.17, 0.05) | 0.2670 |
| HMDB0000562 | creatinine | Carboxylic acids and derivatives | 0.03 (-0.07, 0.14) | 0.5708 | 0.03 (-0.08, 0.14) | 0.5651 |
| HMDB0000641 | glutamine | Carboxylic acids and derivatives | 0 (-0.11, 0.1) | 0.9758 | -0.01 (-0.12, 0.1) | 0.8340 |
| HMDB0000714 | hippurate | Benzene and substituted derivatives | 0.03 (-0.08, 0.14) | 0.5892 | 0.03 (-0.08, 0.14) | 0.5557 |
| HMDB0000767 | pseudouridine | Nucleoside and nucleotide analogues | 0.12 (0.02, 0.23) | 0.0256 | 0.11 (0, 0.21) | 0.0542 |
| HMDB0000929 | tryptophan | Indoles and derivatives | 0.05 (-0.05, 0.16) | 0.3100 | 0.04 (-0.07, 0.14) | 0.4635 |
| HMDB0001008 | biliverdin | Tetrapyrroles and derivatives | -0.12 (-0.23, -0.02) | 0.0249 | -0.1 (-0.21, 0) | 0.0606 |
| HMDB0001046 | cotinine | Pyridines and derivatives | 0.11 (0.01, 0.22) | 0.0387 | 0.12 (0.01, 0.23) | 0.0359 |
| HMDB0001886 | 3-methylxanthine | Imidazopyrimidines | 0.01 (-0.1, 0.12) | 0.8557 | 0.03 (-0.08, 0.14) | 0.5957 |
| HMDB0004824 | N2,N2-dimethylguanosine | Purine nucleosides | 0.08 (-0.03, 0.18) | 0.1593 | 0.06 (-0.04, 0.17) | 0.2520 |
| HMDB0004949 | C16:0 Ceramide (d18:1) | Sphingolipids | 0.12 (0.02, 0.23) | 0.0247 | 0.15 (0.04, 0.25) | 0.0057 |
| HMDB0005923 | N4-acetylcytidine | Pyrimidine nucleosides | 0.09 (-0.02, 0.19) | 0.1133 | 0.07 (-0.03, 0.18) | 0.1835 |
| HMDB0008006* | C34:3 PC | Glycerophospholipids | 0.13 (0.02, 0.23) | 0.0162 | 0.12 (0.02, 0.22) | 0.0257 |
| HMDB0008047* | C38:3 PC | Glycerophospholipids | 0.16 (0.05, 0.26) | 0.0037 | 0.16 (0.05, 0.26) | 0.0040 |
| HMDB0011130 | C18:0 LPE | Glycerophospholipids | -0.02 (-0.13, 0.08) | 0.6731 | -0.01 (-0.12, 0.1) | 0.8480 |
| HMDB0011220* | C36:5 PC plasmalogen-B | Glycerophospholipids | -0.06 (-0.17, 0.05) | 0.2831 | -0.06 (-0.17, 0.05) | 0.2736 |
| *Severe Distress Sample* | | | | | | |
| HMDB0000112 | GABA | Carboxylic acids and derivatives | -0.08 (-0.15, 0) | 0.0506 | -0.07 (-0.15, 0) | 0.0556 |
| HMDB0000167 | threonine | Carboxylic acids and derivatives | 0 (-0.08, 0.08) | 0.9870 | 0 (-0.07, 0.08) | 0.9440 |
| HMDB0000235 | thiamine | Diazines | 0.05 (-0.03, 0.12) | 0.2119 | 0.05 (-0.03, 0.12) | 0.2082 |
| HMDB0000259 | serotonin | Indoles and derivatives | -0.04 (-0.12, 0.03) | 0.2875 | -0.04 (-0.11, 0.04) | 0.3408 |
| HMDB0000562 | creatinine | Carboxylic acids and derivatives | -0.04 (-0.12, 0.03) | 0.2548 | -0.05 (-0.12, 0.03) | 0.2172 |
| HMDB0000641 | glutamine | Carboxylic acids and derivatives | 0.01 (-0.07, 0.08) | 0.8928 | 0.01 (-0.07, 0.08) | 0.8466 |
| HMDB0000714 | hippurate | Benzene and substituted derivatives | -0.06 (-0.14, 0.01) | 0.0887 | -0.07 (-0.14, 0) | 0.0539 |
| HMDB0000767 | pseudouridine | Nucleoside and nucleotide analogues | 0 (-0.07, 0.08) | 0.8977 | -0.01 (-0.08, 0.07) | 0.8771 |
| HMDB0000929 | tryptophan | Indoles and derivatives | -0.05 (-0.12, 0.03) | 0.2297 | -0.04 (-0.12, 0.04) | 0.3006 |
| HMDB0001008 | biliverdin | Tetrapyrroles and derivatives | -0.05 (-0.12, 0.03) | 0.2272 | -0.04 (-0.11, 0.04) | 0.3257 |
| HMDB0001046 | cotinine | Pyridines and derivatives | 0.03 (-0.05, 0.11) | 0.4303 | 0.03 (-0.05, 0.1) | 0.5177 |
| HMDB0001886 | 3-methylxanthine | Imidazopyrimidines | 0.01 (-0.07, 0.08) | 0.8487 | 0 (-0.07, 0.07) | 0.9925 |
| HMDB0004824 | N2,N2-dimethylguanosine | Purine nucleosides | -0.01 (-0.08, 0.06) | 0.7847 | -0.03 (-0.1, 0.04) | 0.4375 |
| HMDB0004949 | C16:0 Ceramide (d18:1) | Sphingolipids | 0.01 (-0.07, 0.08) | 0.8811 | 0.01 (-0.07, 0.08) | 0.8794 |
| HMDB0005923 | N4-acetylcytidine | Pyrimidine nucleosides | 0.04 (-0.04, 0.12) | 0.3116 | 0.03 (-0.04, 0.11) | 0.4008 |
| HMDB0008006* | C34:3 PC | Glycerophospholipids | 0.11 (0.03, 0.18) | 0.0047 | 0.11 (0.03, 0.18) | 0.0046 |
| HMDB0008047* | C38:3 PC | Glycerophospholipids | 0.05 (-0.02, 0.13) | 0.1602 | 0.05 (-0.02, 0.12) | 0.1765 |
| HMDB0011130 | C18:0 LPE | Glycerophospholipids | 0.07 (-0.01, 0.14) | 0.0869 | 0.06 (-0.01, 0.14) | 0.0948 |
| HMDB0011220* | C36:5 PC plasmalogen-B | Glycerophospholipids | 0 (-0.07, 0.07) | 0.9964 | 0.01 (-0.07, 0.08) | 0.8764 |

Supplemental Table S1B. Associations between lifetime PTSD in remission and 19 metabolites previously associated with depression and anxiety included in the metabolite-based distress score, adjusting for base covariates and medical covariates, in the random-effects meta-analysis and within each subsample. Bold values indicate significant associations after correcting for testing 19 metabolites (FDR p<0.05).

| **HMDB ID** | **Metabolite** | **Metabolite Class** | **Minimal**  **β (95% CI)** | **Minimal p-value** | **Model 2**  **β (95% CI)** | **Model 2 p-value** |
| --- | --- | --- | --- | --- | --- | --- |
| *Random-Effects Meta-Analysis* | | | | | | |
| HMDB0000112 | GABA | Carboxylic acids and derivatives | 0.04 (-0.1, 0.19) | 0.5657 | 0.04 (-0.12, 0.19) | 0.6541 |
| HMDB0000167 | threonine | Carboxylic acids and derivatives | -0.29 (-0.8, 0.22) | 0.2652 | -0.31 (-0.82, 0.21) | 0.2420 |
| HMDB0000235 | thiamine | Diazines | 0.07 (-0.08, 0.21) | 0.3634 | 0.06 (-0.09, 0.2) | 0.4406 |
| HMDB0000259 | serotonin | Indoles and derivatives | -0.13 (-0.35, 0.09) | 0.2500 | -0.12 (-0.33, 0.1) | 0.2844 |
| HMDB0000562 | creatinine | Carboxylic acids and derivatives | -0.05 (-0.27, 0.17) | 0.6622 | -0.07 (-0.29, 0.16) | 0.5628 |
| HMDB0000641 | glutamine | Carboxylic acids and derivatives | 0.01 (-0.14, 0.15) | 0.9041 | 0 (-0.14, 0.15) | 0.9661 |
| HMDB0000714 | hippurate | Benzene and substituted derivatives | -0.06 (-0.24, 0.12) | 0.5171 | -0.06 (-0.24, 0.12) | 0.5283 |
| HMDB0000767 | pseudouridine | Nucleoside and nucleotide analogues | 0.03 (-0.11, 0.16) | 0.6929 | 0 (-0.14, 0.13) | 0.9435 |
| HMDB0000929 | tryptophan | Indoles and derivatives | -0.21 (-0.6, 0.18) | 0.2838 | -0.23 (-0.65, 0.18) | 0.2684 |
| HMDB0001008 | biliverdin | Tetrapyrroles and derivatives | 0.07 (-0.07, 0.2) | 0.3180 | 0.08 (-0.06, 0.21) | 0.2759 |
| HMDB0001046 | cotinine | Pyridines and derivatives | 0.07 (-0.12, 0.26) | 0.4890 | 0.04 (-0.18, 0.26) | 0.7286 |
| HMDB0001886 | 3-methylxanthine | Imidazopyrimidines | 0.01 (-0.14, 0.16) | 0.9245 | 0.02 (-0.14, 0.17) | 0.8326 |
| HMDB0004824 | N2,N2-dimethylguanosine | Purine nucleosides | -0.1 (-0.42, 0.22) | 0.5353 | -0.17 (-0.54, 0.21) | 0.3847 |
| HMDB0004949 | C16:0 Ceramide (d18:1) | Sphingolipids | 0 (-0.14, 0.14) | 0.9952 | 0.01 (-0.13, 0.15) | 0.8976 |
| HMDB0005923 | N4-acetylcytidine | Pyrimidine nucleosides | 0.08 (-0.1, 0.26) | 0.3848 | 0.01 (-0.21, 0.24) | 0.8949 |
| HMDB0008006* | C34:3 PC | Glycerophospholipids | -0.04 (-0.36, 0.28) | 0.7979 | -0.02 (-0.44, 0.39) | 0.9103 |
| HMDB0008047* | C38:3 PC | Glycerophospholipids | 0.04 (-0.09, 0.18) | 0.5120 | 0.03 (-0.11, 0.17) | 0.6709 |
| HMDB0011130 | C18:0 LPE | Glycerophospholipids | 0.01 (-0.13, 0.15) | 0.8633 | 0.03 (-0.11, 0.17) | 0.6898 |
| HMDB0011220* | C36:5 PC plasmalogen-B | Glycerophospholipids | -0.02 (-0.16, 0.12) | 0.7511 | -0.02 (-0.16, 0.12) | 0.8249 |
| *NHSII Merged Dataset* | | | | | | |
| HMDB0000112 | GABA | Carboxylic acids and derivatives | 0.07 (-0.08, 0.22) | 0.3668 | 0.07 (-0.09, 0.22) | 0.3882 |
| HMDB0000167 | threonine | Carboxylic acids and derivatives | 0.07 (-0.07, 0.2) | 0.3397 | 0.05 (-0.09, 0.19) | 0.4633 |
| HMDB0000235 | thiamine | Diazines | 0.08 (-0.07, 0.24) | 0.2983 | 0.07 (-0.08, 0.23) | 0.3498 |
| HMDB0000259 | serotonin | Indoles and derivatives | -0.2 (-0.38, -0.02) | 0.0273 | -0.19 (-0.37, -0.02) | 0.0330 |
| HMDB0000562 | creatinine | Carboxylic acids and derivatives | 0 (-0.15, 0.15) | 0.9935 | -0.02 (-0.17, 0.13) | 0.7838 |
| HMDB0000641 | glutamine | Carboxylic acids and derivatives | 0.04 (-0.12, 0.2) | 0.5968 | 0.04 (-0.12, 0.2) | 0.6519 |
| HMDB0000714 | hippurate | Benzene and substituted derivatives | 0 (-0.21, 0.21) | 0.9686 | 0 (-0.21, 0.2) | 0.9769 |
| HMDB0000767 | pseudouridine | Nucleoside and nucleotide analogues | 0.05 (-0.09, 0.19) | 0.5252 | 0.02 (-0.12, 0.16) | 0.8032 |
| HMDB0000929 | tryptophan | Indoles and derivatives | 0.03 (-0.11, 0.17) | 0.6888 | 0.03 (-0.11, 0.17) | 0.6881 |
| HMDB0001008 | biliverdin | Tetrapyrroles and derivatives | 0.09 (-0.05, 0.24) | 0.2015 | 0.1 (-0.05, 0.24) | 0.1970 |
| HMDB0001046 | cotinine | Pyridines and derivatives | 0.12 (-0.04, 0.28) | 0.1351 | 0.11 (-0.05, 0.27) | 0.1647 |
| HMDB0001886 | 3-methylxanthine | Imidazopyrimidines | 0 (-0.16, 0.17) | 0.9835 | 0.01 (-0.15, 0.18) | 0.8743 |
| HMDB0004824 | N2,N2-dimethylguanosine | Purine nucleosides | 0 (-0.15, 0.16) | 0.9579 | -0.02 (-0.18, 0.14) | 0.8406 |
| HMDB0004949 | C16:0 Ceramide (d18:1) | Sphingolipids | -0.03 (-0.17, 0.12) | 0.7369 | -0.02 (-0.17, 0.13) | 0.8096 |
| HMDB0005923 | N4-acetylcytidine | Pyrimidine nucleosides | 0.14 (-0.02, 0.29) | 0.0820 | 0.1 (-0.05, 0.25) | 0.2083 |
| HMDB0008006* | C34:3 PC | Glycerophospholipids | -0.1 (-0.24, 0.04) | 0.1577 | -0.12 (-0.26, 0.02) | 0.0994 |
| HMDB0008047* | C38:3 PC | Glycerophospholipids | 0.06 (-0.08, 0.2) | 0.3863 | 0.05 (-0.1, 0.19) | 0.5406 |
| HMDB0011130 | C18:0 LPE | Glycerophospholipids | 0.01 (-0.13, 0.16) | 0.8428 | 0.03 (-0.12, 0.18) | 0.6825 |
| HMDB0011220* | C36:5 PC plasmalogen-B | Glycerophospholipids | -0.02 (-0.17, 0.12) | 0.7538 | -0.02 (-0.17, 0.13) | 0.7929 |
| *Mind-Body Study* | | | | | | |
| HMDB0000112 | GABA | Carboxylic acids and derivatives | 0.15 (-0.53, 0.84) | 0.6634 | 0.13 (-0.56, 0.82) | 0.7138 |
| HMDB0000167 | threonine | Carboxylic acids and derivatives | -0.86 (-1.53, -0.19) | 0.0122 | -0.88 (-1.56, -0.21) | 0.0111 |
| HMDB0000235 | thiamine | Diazines | 0.42 (-0.25, 1.09) | 0.2188 | 0.37 (-0.3, 1.04) | 0.2797 |
| HMDB0000259 | serotonin | Indoles and derivatives | -0.09 (-0.78, 0.6) | 0.7983 | 0 (-0.68, 0.68) | 0.9946 |
| HMDB0000562 | creatinine | Carboxylic acids and derivatives | 0.21 (-0.46, 0.89) | 0.5374 | 0.23 (-0.45, 0.91) | 0.5081 |
| HMDB0000641 | glutamine | Carboxylic acids and derivatives | -0.51 (-1.19, 0.18) | 0.1501 | -0.53 (-1.22, 0.16) | 0.1351 |
| HMDB0000714 | hippurate | Benzene and substituted derivatives | -0.37 (-1.05, 0.32) | 0.2996 | -0.35 (-1.04, 0.34) | 0.3250 |
| HMDB0000767 | pseudouridine | Nucleoside and nucleotide analogues | 0.13 (-0.55, 0.81) | 0.7121 | 0.04 (-0.63, 0.72) | 0.9043 |
| HMDB0000929 | tryptophan | Indoles and derivatives | -0.68 (-1.35, -0.01) | 0.0465 | -0.73 (-1.4, -0.07) | 0.0323 |
| HMDB0001008 | biliverdin | Tetrapyrroles and derivatives | -0.27 (-0.96, 0.42) | 0.4456 | -0.2 (-0.89, 0.48) | 0.5595 |
| HMDB0001046 | cotinine | Pyridines and derivatives | 0.02 (-0.67, 0.7) | 0.9574 | 0.01 (-0.69, 0.7) | 0.9859 |
| HMDB0001886 | 3-methylxanthine | Imidazopyrimidines | -0.06 (-0.74, 0.62) | 0.8645 | -0.05 (-0.73, 0.62) | 0.8754 |
| HMDB0004824 | N2,N2-dimethylguanosine | Purine nucleosides | 0.16 (-0.53, 0.84) | 0.6537 | 0.08 (-0.6, 0.77) | 0.8169 |
| HMDB0004949 | C16:0 Ceramide (d18:1) | Sphingolipids | 0.34 (-0.33, 1.02) | 0.3207 | 0.38 (-0.28, 1.05) | 0.2605 |
| HMDB0005923 | N4-acetylcytidine | Pyrimidine nucleosides | -0.33 (-1.01, 0.35) | 0.3481 | -0.42 (-1.09, 0.26) | 0.2262 |
| HMDB0008006* | C34:3 PC | Glycerophospholipids | -0.44 (-1.1, 0.23) | 0.1974 | -0.43 (-1.09, 0.23) | 0.2018 |
| HMDB0008047* | C38:3 PC | Glycerophospholipids | -0.41 (-1.08, 0.26) | 0.2336 | -0.43 (-1.1, 0.23) | 0.2050 |
| HMDB0011130 | C18:0 LPE | Glycerophospholipids | -0.43 (-1.12, 0.25) | 0.2152 | -0.42 (-1.11, 0.26) | 0.2290 |
| HMDB0011220* | C36:5 PC plasmalogen-B | Glycerophospholipids | -0.02 (-0.71, 0.67) | 0.9493 | 0.01 (-0.68, 0.69) | 0.9843 |
| *Severe Distress Sample* | | | | | | |
| HMDB0000112 | GABA | Carboxylic acids and derivatives | -0.26 (-0.73, 0.21) | 0.2868 | -0.27 (-0.74, 0.21) | 0.2773 |
| HMDB0000167 | threonine | Carboxylic acids and derivatives | -0.32 (-0.8, 0.16) | 0.1917 | -0.33 (-0.81, 0.14) | 0.1731 |
| HMDB0000235 | thiamine | Diazines | -0.25 (-0.72, 0.22) | 0.2954 | -0.27 (-0.74, 0.21) | 0.2736 |
| HMDB0000259 | serotonin | Indoles and derivatives | 0.14 (-0.33, 0.62) | 0.5530 | 0.13 (-0.35, 0.61) | 0.5922 |
| HMDB0000562 | creatinine | Carboxylic acids and derivatives | -0.38 (-0.85, 0.09) | 0.1117 | -0.41 (-0.88, 0.07) | 0.0965 |
| HMDB0000641 | glutamine | Carboxylic acids and derivatives | -0.05 (-0.51, 0.42) | 0.8458 | -0.04 (-0.51, 0.42) | 0.8569 |
| HMDB0000714 | hippurate | Benzene and substituted derivatives | -0.2 (-0.66, 0.26) | 0.4007 | -0.2 (-0.65, 0.25) | 0.3904 |
| HMDB0000767 | pseudouridine | Nucleoside and nucleotide analogues | -0.23 (-0.7, 0.24) | 0.3346 | -0.27 (-0.73, 0.2) | 0.2583 |
| HMDB0000929 | tryptophan | Indoles and derivatives | -0.29 (-0.77, 0.19) | 0.2360 | -0.29 (-0.77, 0.2) | 0.2439 |
| HMDB0001008 | biliverdin | Tetrapyrroles and derivatives | -0.04 (-0.51, 0.43) | 0.8596 | 0 (-0.48, 0.47) | 0.9901 |
| HMDB0001046 | cotinine | Pyridines and derivatives | -0.21 (-0.68, 0.27) | 0.4012 | -0.25 (-0.73, 0.24) | 0.3199 |
| HMDB0001886 | 3-methylxanthine | Imidazopyrimidines | 0.08 (-0.38, 0.54) | 0.7293 | 0.07 (-0.39, 0.54) | 0.7576 |
| HMDB0004824 | N2,N2-dimethylguanosine | Purine nucleosides | -0.47 (-0.93, -0.01) | 0.0483 | -0.56 (-1, -0.12) | 0.0136 |
| HMDB0004949 | C16:0 Ceramide (d18:1) | Sphingolipids | 0.09 (-0.38, 0.55) | 0.7203 | 0.09 (-0.38, 0.57) | 0.6967 |
| HMDB0005923 | N4-acetylcytidine | Pyrimidine nucleosides | -0.04 (-0.51, 0.44) | 0.8792 | -0.08 (-0.55, 0.39) | 0.7483 |
| HMDB0008006* | C34:3 PC | Glycerophospholipids | 0.33 (-0.14, 0.8) | 0.1688 | 0.41 (-0.05, 0.88) | 0.0823 |
| HMDB0008047* | C38:3 PC | Glycerophospholipids | 0.07 (-0.4, 0.53) | 0.7809 | 0.09 (-0.38, 0.56) | 0.7028 |
| HMDB0011130 | C18:0 LPE | Glycerophospholipids | 0.19 (-0.28, 0.66) | 0.4237 | 0.22 (-0.26, 0.69) | 0.3731 |
| HMDB0011220* | C36:5 PC plasmalogen-B | Glycerophospholipids | -0.01 (-0.48, 0.46) | 0.9691 | 0.01 (-0.45, 0.48) | 0.9499 |

Supplemental Table S1C. Associations between trauma exposure and 19 metabolites previously associated with depression and anxiety included in the metabolite-based distress score, adjusting for base covariates and medical covariates, in the random-effects meta-analysis and within each subsample. Bold values indicate significant associations after correcting for testing 19 metabolites (FDR p<0.05).

| **HMDB ID** | **Metabolite** | **Metabolite Class** | **Minimal**  **β (95% CI)** | **Minimal p-value** | **Model 2**  **β (95% CI)** | **Model 2 p-value** |
| --- | --- | --- | --- | --- | --- | --- |
| *Random-Effects Meta-Analysis* | | | | | | |
| HMDB0000112 | GABA | Carboxylic acids and derivatives | 0.1 (-0.13, 0.34) | 0.3910 | 0.11 (-0.14, 0.35) | 0.3932 |
| HMDB0000167 | threonine | Carboxylic acids and derivatives | -0.05 (-0.14, 0.04) | 0.3073 | -0.05 (-0.14, 0.04) | 0.2751 |
| HMDB0000235 | thiamine | Diazines | -0.06 (-0.23, 0.11) | 0.4836 | -0.05 (-0.21, 0.1) | 0.4849 |
| HMDB0000259 | serotonin | Indoles and derivatives | -0.04 (-0.13, 0.05) | 0.3464 | -0.05 (-0.14, 0.04) | 0.2926 |
| HMDB0000562 | creatinine | Carboxylic acids and derivatives | 0.14 (-0.3, 0.57) | 0.5421 | 0.14 (-0.32, 0.59) | 0.5505 |
| HMDB0000641 | glutamine | Carboxylic acids and derivatives | -0.04 (-0.13, 0.05) | 0.4116 | -0.04 (-0.13, 0.05) | 0.3833 |
| HMDB0000714 | hippurate | Benzene and substituted derivatives | 0 (-0.19, 0.18) | 0.9597 | 0.01 (-0.2, 0.23) | 0.8997 |
| HMDB0000767 | pseudouridine | Nucleoside and nucleotide analogues | 0.05 (-0.05, 0.14) | 0.3353 | 0.05 (-0.04, 0.14) | 0.2707 |
| HMDB0000929 | tryptophan | Indoles and derivatives | 0.03 (-0.06, 0.12) | 0.4808 | 0.03 (-0.06, 0.12) | 0.5329 |
| HMDB0001008 | biliverdin | Tetrapyrroles and derivatives | 0.01 (-0.08, 0.11) | 0.7590 | 0.01 (-0.08, 0.1) | 0.7937 |
| HMDB0001046 | cotinine | Pyridines and derivatives | 0.03 (-0.06, 0.12) | 0.4581 | 0.04 (-0.05, 0.13) | 0.4315 |
| HMDB0001886 | 3-methylxanthine | Imidazopyrimidines | 0.05 (-0.05, 0.14) | 0.3540 | 0.04 (-0.06, 0.14) | 0.4198 |
| HMDB0004824 | N2,N2-dimethylguanosine | Purine nucleosides | 0.01 (-0.38, 0.4) | 0.9527 | 0.05 (-0.41, 0.5) | 0.8416 |
| HMDB0004949 | C16:0 Ceramide (d18:1) | Sphingolipids | -0.05 (-0.3, 0.2) | 0.6798 | -0.07 (-0.33, 0.2) | 0.6227 |
| HMDB0005923 | N4-acetylcytidine | Pyrimidine nucleosides | 0.03 (-0.06, 0.12) | 0.5001 | 0.04 (-0.05, 0.13) | 0.3773 |
| HMDB0008006* | C34:3 PC | Glycerophospholipids | -0.13 (-0.52, 0.26) | 0.5128 | -0.12 (-0.52, 0.27) | 0.5321 |
| HMDB0008047* | C38:3 PC | Glycerophospholipids | -0.11 (-0.43, 0.2) | 0.4785 | -0.11 (-0.43, 0.22) | 0.5120 |
| HMDB0011130 | C18:0 LPE | Glycerophospholipids | -0.14 (-0.55, 0.28) | 0.5170 | -0.14 (-0.55, 0.26) | 0.4817 |
| HMDB0011220* | C36:5 PC plasmalogen-B | Glycerophospholipids | -0.03 (-0.13, 0.06) | 0.4846 | -0.03 (-0.13, 0.06) | 0.4892 |
| *NHSII Merged Dataset* | | | | | | |
| HMDB0000112 | GABA | Carboxylic acids and derivatives | -0.03 (-0.13, 0.07) | 0.5288 | -0.04 (-0.14, 0.06) | 0.4763 |
| HMDB0000167 | threonine | Carboxylic acids and derivatives | -0.06 (-0.15, 0.04) | 0.2470 | -0.06 (-0.15, 0.03) | 0.2103 |
| HMDB0000235 | thiamine | Diazines | -0.02 (-0.11, 0.07) | 0.6997 | -0.02 (-0.11, 0.07) | 0.6810 |
| HMDB0000259 | serotonin | Indoles and derivatives | -0.05 (-0.14, 0.04) | 0.2980 | -0.05 (-0.15, 0.04) | 0.2736 |
| HMDB0000562 | creatinine | Carboxylic acids and derivatives | -0.09 (-0.18, 0) | 0.0557 | -0.09 (-0.19, 0) | 0.0438 |
| HMDB0000641 | glutamine | Carboxylic acids and derivatives | -0.03 (-0.12, 0.07) | 0.6177 | -0.03 (-0.13, 0.07) | 0.5832 |
| HMDB0000714 | hippurate | Benzene and substituted derivatives | -0.08 (-0.2, 0.04) | 0.2127 | -0.08 (-0.2, 0.04) | 0.2091 |
| HMDB0000767 | pseudouridine | Nucleoside and nucleotide analogues | 0.06 (-0.04, 0.15) | 0.2671 | 0.06 (-0.04, 0.16) | 0.2432 |
| HMDB0000929 | tryptophan | Indoles and derivatives | 0.03 (-0.06, 0.13) | 0.5009 | 0.03 (-0.07, 0.12) | 0.5537 |
| HMDB0001008 | biliverdin | Tetrapyrroles and derivatives | 0 (-0.1, 0.09) | 0.9425 | 0 (-0.1, 0.1) | 0.9847 |
| HMDB0001046 | cotinine | Pyridines and derivatives | 0.02 (-0.07, 0.11) | 0.6734 | 0.02 (-0.07, 0.12) | 0.6393 |
| HMDB0001886 | 3-methylxanthine | Imidazopyrimidines | 0.04 (-0.06, 0.15) | 0.4185 | 0.04 (-0.07, 0.14) | 0.5003 |
| HMDB0004824 | N2,N2-dimethylguanosine | Purine nucleosides | 0 (-0.1, 0.09) | 0.9215 | -0.01 (-0.1, 0.09) | 0.8852 |
| HMDB0004949 | C16:0 Ceramide (d18:1) | Sphingolipids | 0.08 (-0.02, 0.18) | 0.1126 | 0.09 (-0.01, 0.19) | 0.0745 |
| HMDB0005923 | N4-acetylcytidine | Pyrimidine nucleosides | 0.04 (-0.05, 0.14) | 0.4016 | 0.05 (-0.05, 0.15) | 0.3096 |
| HMDB0008006* | C34:3 PC | Glycerophospholipids | -0.06 (-0.15, 0.04) | 0.2328 | -0.06 (-0.15, 0.03) | 0.2061 |
| HMDB0008047* | C38:3 PC | Glycerophospholipids | 0.08 (-0.02, 0.17) | 0.1212 | 0.09 (0, 0.18) | 0.0630 |
| HMDB0011130 | C18:0 LPE | Glycerophospholipids | 0.01 (-0.09, 0.1) | 0.8946 | 0.01 (-0.09, 0.1) | 0.9168 |
| HMDB0011220* | C36:5 PC plasmalogen-B | Glycerophospholipids | -0.01 (-0.11, 0.08) | 0.7765 | -0.01 (-0.11, 0.09) | 0.7849 |
| *Mind-Body Study* | | | | | | |
| HMDB0000112 | GABA | Carboxylic acids and derivatives | 0.2 (-0.22, 0.62) | 0.3595 | 0.21 (-0.21, 0.64) | 0.3317 |
| HMDB0000167 | threonine | Carboxylic acids and derivatives | -0.2 (-0.61, 0.21) | 0.3285 | -0.18 (-0.59, 0.24) | 0.4006 |
| HMDB0000235 | thiamine | Diazines | -0.4 (-0.81, 0.01) | 0.0565 | -0.39 (-0.81, 0.02) | 0.0622 |
| HMDB0000259 | serotonin | Indoles and derivatives | 0.22 (-0.2, 0.64) | 0.3060 | 0.19 (-0.24, 0.61) | 0.3890 |
| HMDB0000562 | creatinine | Carboxylic acids and derivatives | -0.06 (-0.48, 0.35) | 0.7687 | -0.07 (-0.49, 0.35) | 0.7350 |
| HMDB0000641 | glutamine | Carboxylic acids and derivatives | -0.14 (-0.56, 0.28) | 0.5087 | -0.13 (-0.56, 0.3) | 0.5478 |
| HMDB0000714 | hippurate | Benzene and substituted derivatives | -0.04 (-0.47, 0.38) | 0.8392 | -0.06 (-0.49, 0.37) | 0.7826 |
| HMDB0000767 | pseudouridine | Nucleoside and nucleotide analogues | -0.3 (-0.72, 0.12) | 0.1688 | -0.29 (-0.7, 0.13) | 0.1788 |
| HMDB0000929 | tryptophan | Indoles and derivatives | 0.03 (-0.38, 0.44) | 0.8914 | 0.06 (-0.35, 0.47) | 0.7623 |
| HMDB0001008 | biliverdin | Tetrapyrroles and derivatives | 0.07 (-0.36, 0.49) | 0.7578 | 0.03 (-0.39, 0.46) | 0.8723 |
| HMDB0001046 | cotinine | Pyridines and derivatives | 0.22 (-0.2, 0.64) | 0.3139 | 0.19 (-0.23, 0.62) | 0.3754 |
| HMDB0001886 | 3-methylxanthine | Imidazopyrimidines | 0.38 (-0.04, 0.8) | 0.0788 | 0.33 (-0.09, 0.75) | 0.1239 |
| HMDB0004824 | N2,N2-dimethylguanosine | Purine nucleosides | -0.38 (-0.8, 0.04) | 0.0787 | -0.37 (-0.79, 0.05) | 0.0892 |
| HMDB0004949 | C16:0 Ceramide (d18:1) | Sphingolipids | -0.03 (-0.44, 0.38) | 0.8879 | -0.09 (-0.5, 0.32) | 0.6591 |
| HMDB0005923 | N4-acetylcytidine | Pyrimidine nucleosides | -0.05 (-0.47, 0.37) | 0.8069 | -0.05 (-0.46, 0.37) | 0.8177 |
| HMDB0008006* | C34:3 PC | Glycerophospholipids | 0.2 (-0.2, 0.61) | 0.3278 | 0.22 (-0.19, 0.63) | 0.2909 |
| HMDB0008047* | C38:3 PC | Glycerophospholipids | -0.07 (-0.48, 0.34) | 0.7345 | -0.06 (-0.47, 0.35) | 0.7677 |
| HMDB0011130 | C18:0 LPE | Glycerophospholipids | 0.15 (-0.27, 0.57) | 0.4930 | 0.12 (-0.3, 0.54) | 0.5829 |
| HMDB0011220* | C36:5 PC plasmalogen-B | Glycerophospholipids | -0.17 (-0.59, 0.25) | 0.4315 | -0.15 (-0.57, 0.27) | 0.4881 |
| *Severe Distress Sample* | | | | | | |
| HMDB0000112 | GABA | Carboxylic acids and derivatives | 0.33 (-0.04, 0.69) | 0.0844 | 0.33 (-0.05, 0.7) | 0.0882 |
| HMDB0000167 | threonine | Carboxylic acids and derivatives | 0.23 (-0.15, 0.6) | 0.2350 | 0.22 (-0.15, 0.59) | 0.2513 |
| HMDB0000235 | thiamine | Diazines | 0.05 (-0.32, 0.41) | 0.7905 | 0.05 (-0.32, 0.42) | 0.7941 |
| HMDB0000259 | serotonin | Indoles and derivatives | -0.13 (-0.5, 0.23) | 0.4763 | -0.16 (-0.53, 0.21) | 0.4104 |
| HMDB0000562 | creatinine | Carboxylic acids and derivatives | **0.6 (0.24, 0.97)** | **0.0014** | **0.63 (0.26, 1)** | **0.0011** |
| HMDB0000641 | glutamine | Carboxylic acids and derivatives | -0.15 (-0.51, 0.21) | 0.4241 | -0.16 (-0.52, 0.2) | 0.3834 |
| HMDB0000714 | hippurate | Benzene and substituted derivatives | 0.25 (-0.11, 0.6) | 0.1801 | 0.28 (-0.07, 0.63) | 0.1182 |
| HMDB0000767 | pseudouridine | Nucleoside and nucleotide analogues | 0.16 (-0.2, 0.52) | 0.3851 | 0.22 (-0.14, 0.58) | 0.2377 |
| HMDB0000929 | tryptophan | Indoles and derivatives | 0.03 (-0.34, 0.4) | 0.8729 | 0 (-0.38, 0.37) | 0.9895 |
| HMDB0001008 | biliverdin | Tetrapyrroles and derivatives | 0.23 (-0.14, 0.59) | 0.2235 | 0.18 (-0.19, 0.55) | 0.3327 |
| HMDB0001046 | cotinine | Pyridines and derivatives | 0.1 (-0.27, 0.47) | 0.5941 | 0.13 (-0.25, 0.5) | 0.5158 |
| HMDB0001886 | 3-methylxanthine | Imidazopyrimidines | -0.16 (-0.52, 0.2) | 0.3769 | -0.13 (-0.49, 0.24) | 0.4927 |
| HMDB0004824 | N2,N2-dimethylguanosine | Purine nucleosides | 0.39 (0.03, 0.75) | 0.0345 | **0.49 (0.15, 0.83)** | **0.0058** |
| HMDB0004949 | C16:0 Ceramide (d18:1) | Sphingolipids | -0.34 (-0.7, 0.03) | 0.0703 | -0.34 (-0.71, 0.03) | 0.0728 |
| HMDB0005923 | N4-acetylcytidine | Pyrimidine nucleosides | -0.05 (-0.42, 0.32) | 0.7879 | -0.02 (-0.39, 0.35) | 0.9167 |
| HMDB0008006* | C34:3 PC | Glycerophospholipids | **-0.54 (-0.9, -0.18)** | **0.0041** | **-0.53 (-0.89, -0.17)** | **0.0042** |
| HMDB0008047* | C38:3 PC | Glycerophospholipids | -0.45 (-0.81, -0.08) | 0.0167 | -0.45 (-0.81, -0.09) | 0.0161 |
| HMDB0011130 | C18:0 LPE | Glycerophospholipids | **-0.58 (-0.95, -0.22)** | **0.0019** | **-0.59 (-0.95, -0.22)** | **0.0022** |
| HMDB0011220* | C36:5 PC plasmalogen-B | Glycerophospholipids | -0.19 (-0.56, 0.17) | 0.3063 | -0.2 (-0.56, 0.16) | 0.2789 |

Supplemental Table S2. Associations between the three exposure variables (persistent PTSD symptoms, lifetime PTSD in remission, and trauma exposure) and all 339 metabolites examined in the agnostic analysis. Results from all three models with incremental covariate adjustments are presented. Highlighted values indicate significant associations after testing 339 metabolites (FDR p<0.05).

*See the excel file attached.*

| Supplementary Table S3. Network centrality measures for metabolites significantly associated with persistent PTSD symptoms in the minimal model and had significant edges in the differential network analysis (FDR p<0.05). For hub centrality and closeness centrality, bold face indicates a metabolite was in the top five. For betweenness centrality, bold face indicates a metabolite had nonzero betweenness. Closeness centrality is shown in units of 10^-4^. | | | | |
| --- | --- | --- | --- | --- |
| Metabolite | Degree | Hub Score | Betweenness | Closeness |
| C50:4 TAG | 14 | **1.00** | **94** | **19.79** |
| C50:3 TAG | 14 | 0.00 | 0 | 14.25 |
| C50:5 TAG | 12 | 0.00 | 0 | 14.54 |
| C36:1 PE | 11 | **0.37** | 0 | 14.25 |
| C52:1 TAG | 9 | **0.42** | **15** | **15.42** |
| C34:2 PC | 8 | 0.00 | **18** | **17.62** |
| C22:0 Ceramide (d18:1) | 6 | **0.38** | 0 | **15.20** |
| C10:2 carnitine | 6 | 0.00 | 0 | 11.44 |
| C54:1 TAG | 5 | 0.00 | 0 | **16.21** |
| aminoisobutyric acid | 4 | 0.22 | 0 | 12.58 |
| C55:2 TAG | 4 | 0.09 | 0 | 8.69 |
| homoarginine | 4 | 0.00 | 0 | 12.71 |
| C36:2 PE | 3 | **0.24** | 0 | 13.19 |
| C38:4 PE | 3 | 0.00 | 0 | 13.71 |
| C34:3 PC | 2 | 0.23 | 0 | 13.86 |
| C36:2 DAG | 2 | 0.00 | 0 | 5.45 |
| C34:1 DAG | 1 | 0.01 | 0 | 3.27 |

Supplementary Table S4. Weights used to construct the metabolite-based distress score. Arachidonate was unavailable in our subsamples.

| **Metabolite** | **HMDB ID** | **MDS weight applied** |
| --- | --- | --- |
| serotonin | HMDB0000259 | -0.59 |
| N2,N2-dimethylguanosine | HMDB0004824 | 0.57 |
| threonine | HMDB0000167 | -0.44 |
| hippurate | HMDB0000714 | -0.43 |
| biliverdin | HMDB0001008 | -0.42 |
| C34:3 PC | HMDB0008006 | -0.4 |
| glutamine | HMDB0000641 | 0.37 |
| cotinine | HMDB0001046 | 0.37 |
| creatinine | HMDB0000562 | -0.36 |
| arachidonate | HMDB0001043 | -0.33 |
| 3-methylxanthine | HMDB0001886 | 0.33 |
| C38:3 PC | HMDB0008047 | 0.31 |
| tryptophan | HMDB0000929 | -0.25 |
| GABA | HMDB0000112 | -0.15 |
| C16:0 Ceramide (d18:1) | HMDB0004949 | 0.13 |
| N4-acetylcytidine | HMDB0005923 | -0.12 |
| C36:5 PC plasmalogen-B | HMDB0011220 | -0.11 |
| C18:0 LPE | HMDB0011130 | 0.11 |
| pseudouridine | HMDB0000767 | 0.05 |
| thiamine | HMDB0000235 | 0.05 |

Supplemental Figure S1. Study timeline.

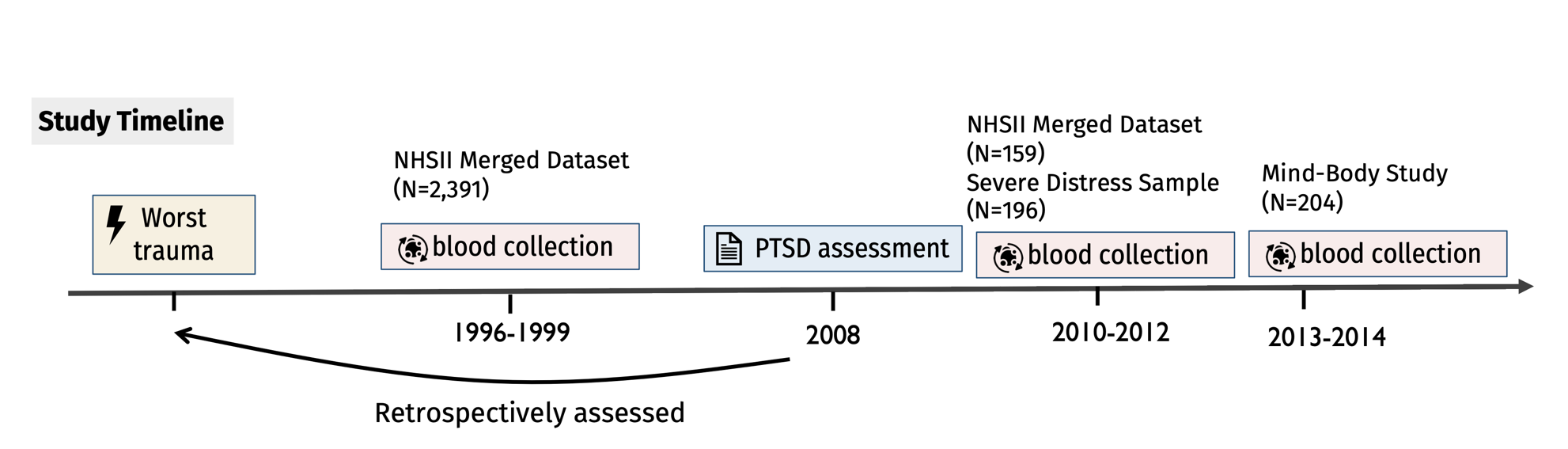

Supplemental Figure S2. Metabolite set enrichment analysis of HMDB metabolite class, comparing associations with persistent PTSD symptoms estimated in the current study and associations with depression/anxiety in the validation dataset reported by Shutta et al. (2021).

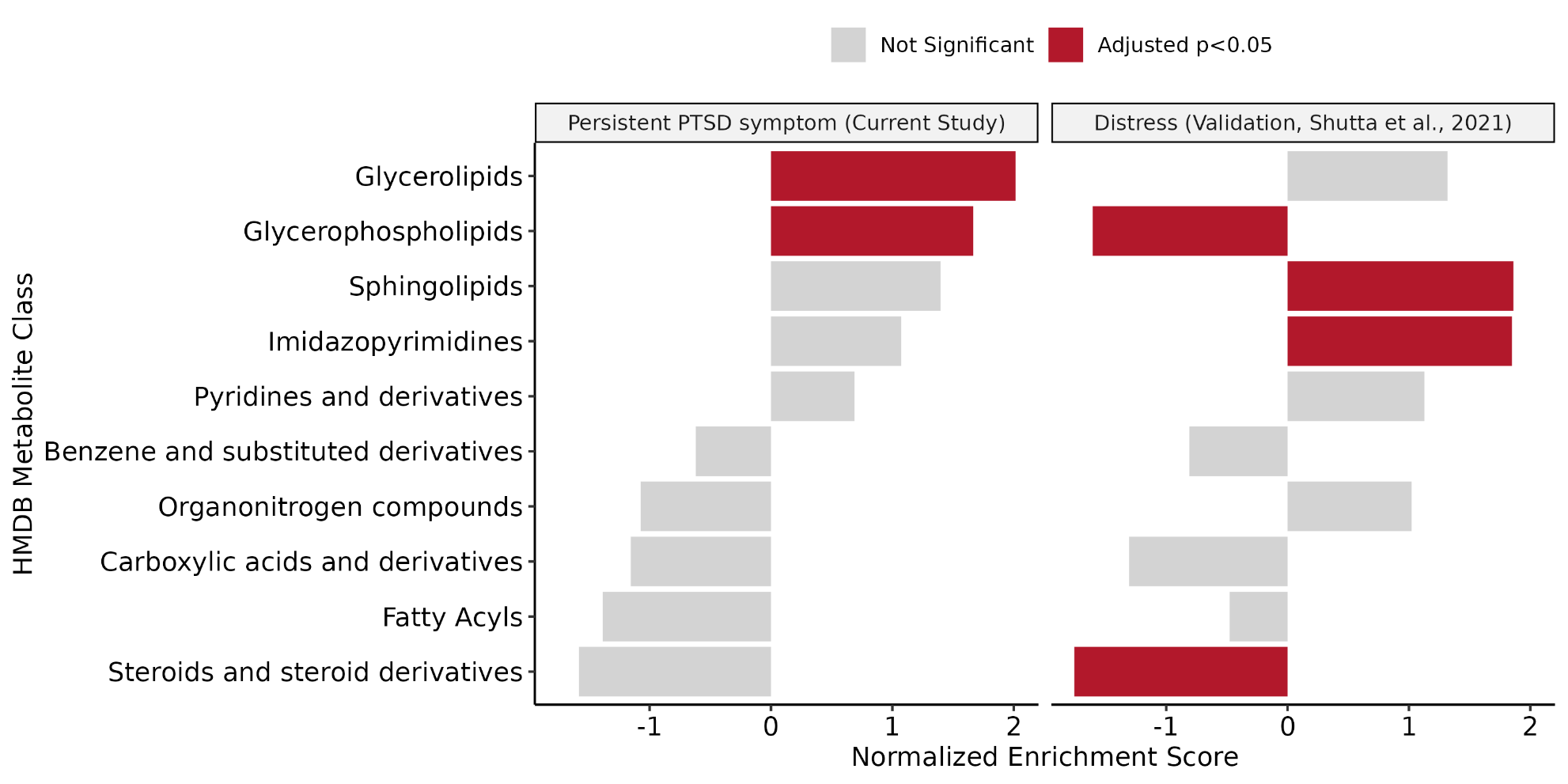

Supplemental Figure S3. Comparisons between results from the primary analysis and sensitivity analysis restricted to controls in the case-control sub-studies within the NHSII merged dataset, i.e., individuals who never developed the physical condition endpoint in each sub-study (N=1944).

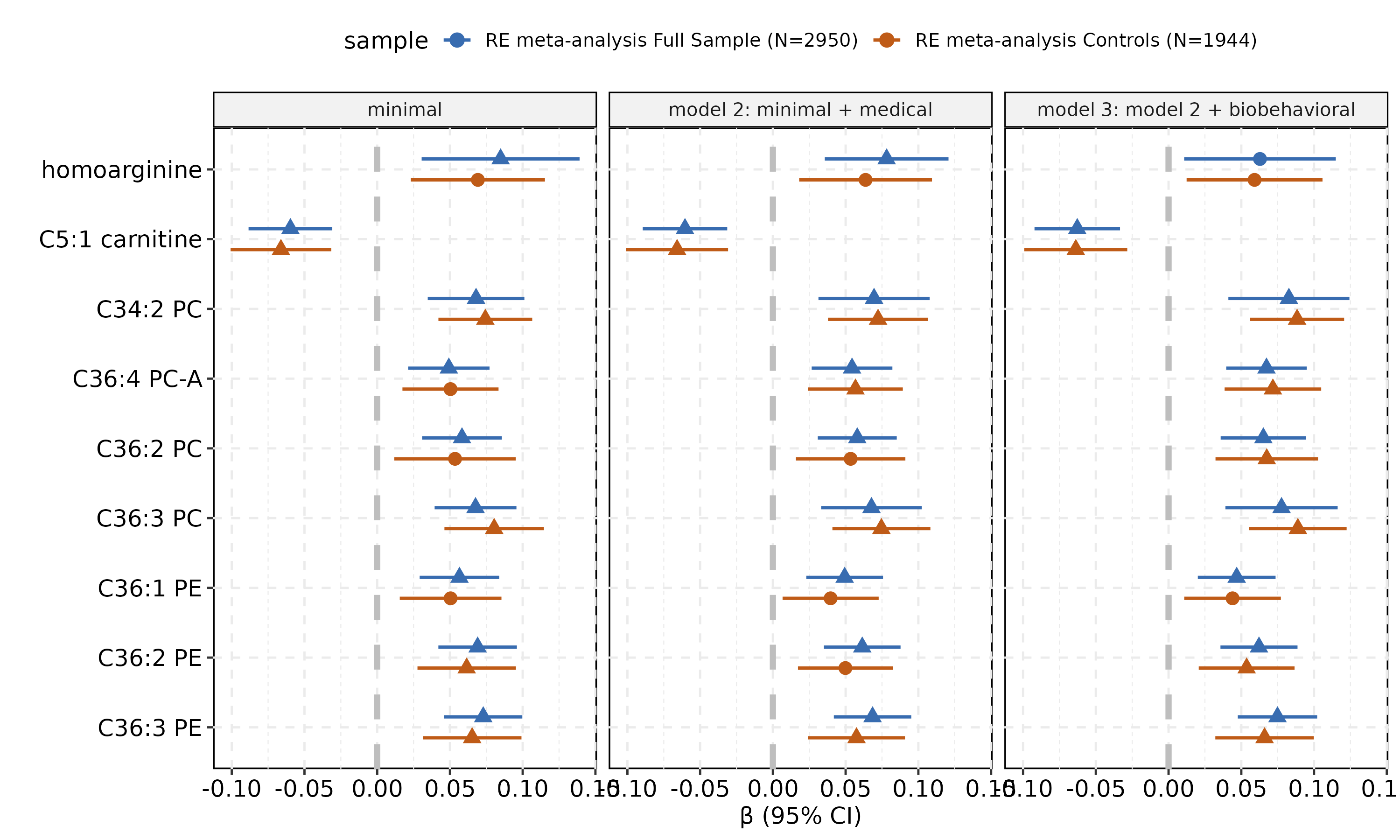
